# Prevalence of hereditary tubulointerstitial kidney diseases in the German Chronic Kidney Disease study

**DOI:** 10.1101/2021.09.29.21264100

**Authors:** Bernt Popp, Arif B. Ekici, Karl X. Knaup, Karen Schneider, Steffen Uebe, Jonghun Park, Vineet Bafna, Heike Meiselbach, Kai-Uwe Eckardt, Mario Schiffer, André Reis, Cornelia Kraus, Michael Wiesener

## Abstract

Exome sequencing (ES) studies in chronic kidney disease (CKD) cohorts could identify pathogenic variants in ∼10% of patients. This implies underdiagnosis of hereditary CKD. Tubulointerstitial kidney diseases, showing no typical clinical/histologic finding but tubulointerstitial fibrosis, are particularly difficult to diagnose.

We used a custom designed targeted panel (29 genes) and *MUC1*-SNaPshot to sequence 271 DNA samples, selected by clinical criteria from 5,217 individuals in the German Chronic Kidney Disease (GCKD) cohort.

We identified 33 pathogenic small variants. Of these 27 (81.8%) were in COL4-genes, the largest group being 15 *COL4A5*-variants with nine unrelated individuals carrying c.1871G>A, p.(Gly624Asp). We found three cysteine variants in *UMOD*, a novel missense, and a novel splice variant in *HNF1B* and the homoplastic *MTTF* variant m.616T>C. Copy-number analysis identified a heterozygous *COL4A5-*deletion, and a *HNF1B*-duplication/-deletion, respectively. Overall, pathogenic variants were present in 12.5% (34/271) and variants of unknown significance in 9.6% (26/271) of selected individuals. Bioinformatic predictions paired with gold standard diagnostics for *MUC1* (SNaPshot) could not identify the typical cytosine duplication (“c.428dupC”) in any individual, implying that ADTKD-*MUC1* is rare.

Our study shows that >10% of individuals with certain clinical features carry disease variants in genes associated with tubulointerstitial kidney diseases. COL4-genes constitute the largest fraction, implying they are overlooked using clinical Alport-syndrome criteria. We also identified variants easily missed by some ES pipelines. Finally, our results indicate that the filtering criteria applied enrich for an underlying genetic disorder.

**SIGNIFICANCE STATEMENT:** CKD affects >10% of the global population and recent studies imply that a considerable portion can be attributed to monogenic diseases, which are likely underappreciated in the clinical routine. Tubulointerstitial kidney diseases are a particularly difficult group of hereditary kidney diseases to diagnose both clinically and genetically. To investigate the prevalence of these disorders in a large CKD cohort we established a set of clinical criteria and designed a custom panel sequencing pipeline. Based on the diagnostic yield of 12.5%, we recommend an algorithm to clinically select and genetically evaluate patients with increased risk for a hereditary tubulointerstitial kidney disease.

## INTRODUCTION

Genetic kidney diseases are underdiagnosed, yet recent data imply that they are much more frequent than the clinical perception. The complexity amongst hereditary kidney diseases is high, with more than 200 diseases and considerably more candidate genes being associated.^1,2^ A systematic approach using exome sequencing (ES) in a cohort of more than 3.000 patients with chronic kidney disease (CKD) has recently yielded diagnostic variants in almost 10% of patients.^3^ Further studies with similar results have been published using ES on different patient cohorts, either population based or selected by specific disease entities. In these studies the diagnostic yield has been reported between 7% and 40% depending on population characteristics and selection criteria (e.g. pediatric vs. adult, syndromic vs. isolated, familial vs. simplex)^4^. The number of hereditary kidney diseases is likely higher, since less clear genetic variants and genes not reliably associated with CKD have been excluded and complex genomic regions (such as repeat sequences) and diseases caused by copy number variants (CNVs) may be difficult to identify by ES^5^. Furthermore, mitochondrial diseases are regularly missed since the mitochondrial genome (mtDNA) is not targeted in typical ES designs. Therefore, the true prevalence of genetic diseases among patients with CKD remains ambiguous to date.

A particularly difficult group of hereditary kidney diseases to diagnose are tubulointerstitial kidney diseases. These diseases cannot be recognized by any typical clinical or histopathological signs. They are characterized merely by progressive CKD and secondary features such as hypertension, as well as tubulointerstitial fibrosis in the kidney biopsy. Specific hereditary diseases with a fibrotic, tubulointerstitial phenotype primarily affecting the adult are autosomal dominant tubulointerstitial kidney diseases (ADTKD)^6,7^ and mitochondrially inherited tubulointerstitial kidney diseases (MITKD)^8^. Furthermore, the heterogeneous group of nephronophthisis (NPHP; considered pediatric^9^) would also meet these criteria. large investigative adult CKD cohorts have shown an unexpected highL prevalence of Collagen-4 (COL4) diseases, possibly also in patients not predicted as being affected from a primary glomerular disease^3^. Thus, searching for hereditary diseases with a tubulointerstitial phenotype should reasonably include genes associated with ADTKD, MITKD, NPHP and COL4-diseases. Some of these disease entities will not be detected by standard next generation sequencing (NGS) techniques, which is particularly true for ADTKD-*MUC1*,^10,11^ ADTKD-*HNF1B* where up to 50% of mutations consist of CNVs^12^ and MITKD^8^. Therefore, a comprehensive search for tubulointerstitial diseases should include technological options to detect these diseases.

To investigate the prevalence of these disorders in a large CKD cohort we established a set of clinical criteria to select individuals with increased risk for tubulointerstitial diseases from the >5.000 adult individuals previously recruited into the German Chronic Kidney Disease (GCKD)^13,14^ cohort. To ameliorate some of the diagnostic gaps of ES and enable rapid and high quality sequencing of our cohort, we designed a custom sequencing panel paired with a bioinformatic pipeline enabling analysis of copy number, mitochondrial variants and the *MUC1*-VNTR. Selected samples were subject to sequencing which was supplemented with gold-standard *MUC1*-dupC diagnostics by SNaPshot^11^.

Based on the diagnostic yield of our study and in comparison with published screenings, we recommend an algorithm to select patients with increased risk for a hereditary tubulointerstitial kidney disease for genetic diagnostics and propose sequencing assays and accompanying analysis pipelines for rare kidney diseases.

## MATERIALS AND METHODS

### Ethics and study cohort

This study adheres to the principles set out in the Declaration of Helsinki. The probands included in our study were filtered using the database of the German Chronic Kidney Disease (GCKD) cohort which enrolled 5,217 individuals. The GCKD study is registered as a national clinical study (DRKS 00003971). It was approved by local ethics review boards of all participating institutions^14^. The Ethical Committee of the Friedrich-Alexander University (FAU) Erlangen-Nürnberg approved the study protocol (“Prospektive Beobachtungsstudie-German Chronic Kidney Disease (GCKD)-Studie”; Re.-Nr. 3831; decision date: 3.7.2008) representing the GCKD lead center at FAU.

The GCKD database was filtered using nine annotated categories (“nephrosclerosis”, “gout”, “IgA nephropathy”, “chronic glomerulonephritis”, “analgesic nephropathy”, “interstitial nephritis”, “hereditary disorders”, “others”, “unknown”) considering the individual’s age (cutoff <= 50 years, except “IgA nephropathy”, “chronic glomerulonephritis”, “analgesic nephropathy” with <= 40 years and “hereditary disorders” with no age cutoff) as presumed leading CKD etiology. We excluded all individuals with known postrenal or primary glomerular disease etiology, known systemic disease, known status after acute kidney injury, polycystic kidneys and those with single kidneys. Biobank DNA samples were subsequently picked and analyzed for quality. All filtering and quality control steps are depicted in Figure 1A.

**Figure 1.**
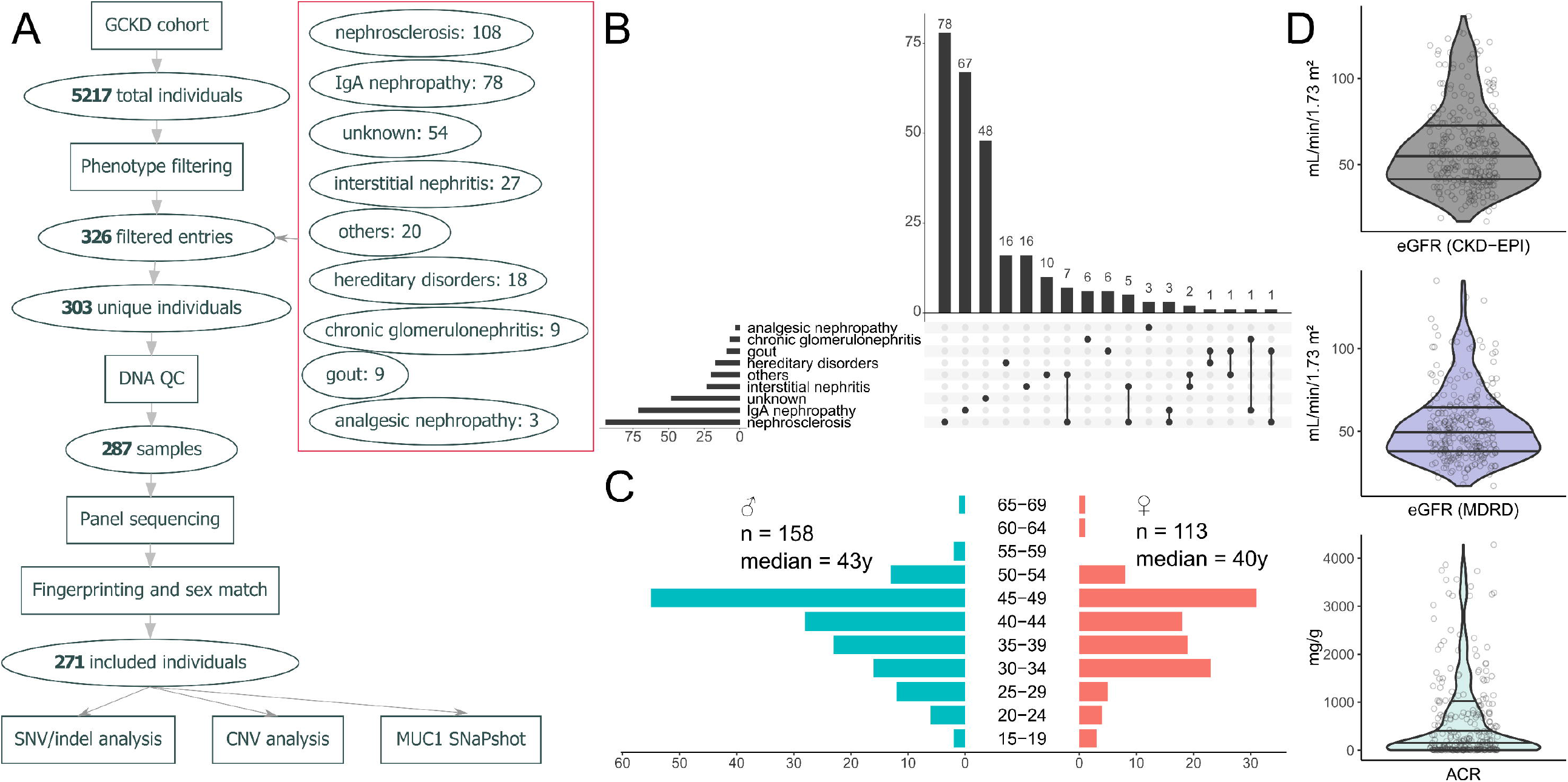
Filtering approach and cohort characteristics. (**A**) Workflow used to filter individuals from the GCKD and subsequent quality control steps to ensure DNA integrity and sample identity. Clinical category based filtering resulted in 326 entries which corresponded to 303 unique individuals (5.8% of the whole GCKD cohort). Of these 271 (89.4%) passed all quality control steps and were included in the final analyses. (**B**) Upset plot showing the distribution and overlap of the nine clinical criteria used to filter the study cohort from the GCKD cohort. (**C**) Age distribution by sex in the final cohort. The y-axis depicts age classes 5-year intervals. The x-axis shows the number of individuals, with females on the right (red) and males on the left (blue) side. Age is reported at inclusion into the GCKD study. (**D**) Distribution of different kidney function parameters at GCKD study inclusion: Top (dark grey) eGFR by CKD-EPI, Middle (blue) eGFR by MDRD, Bottom (light grey) Albumin-Creatinine Ratio (ACR).

### Custom targeted panel design

To design a custom panel covering genes associated with tubulointerstitial kidney disease phenotype, we screened the literature for publications on ADTKD and associated genes, which resulted in the inclusion of the five genes *MUC1, UMOD, REN, HNF1B, SEC61A1* ^10,15–18^. To investigate potential bioinformatic approaches of detecting *MUC1* frameshift variants typically located in the VNTR between exon 2 and 3, custom probes covering this region were included and three individuals from families with a *MUC1-dupC* variant confirmed previously by SNaPshot ^11^ and long-read sequencing ^19^ were sequenced as controls. We also included three recently published differential diagnoses for ADTKD (genes *DNAJB1, GATM, PARN*) and 17 nephronophthisis genes. As individuals with ADTKD can have moderate hematuria or proteinuria we also included the three Collagen 4 genes (*COL4A3, COL4A4, COL4A5*). Due to the association of tubulointerstitial kidney disease with mitochondrial variants, we added capture probes covering the complete mitochondrial genome. Six gene loci on the X-chromosome (sex computation from coverage) and 24 single nucleotide polymorphism (SNP; genomic fingerprinting) markers were added for quality control. Full details on the panel design can be found in File S2.

### Bioinformatic pipeline

Resulting sequence files in BCL format were demultiplexed using bcl2fastq version v1.8.4 (Illumina, Inc., San Diego, CA, USA). Resulting paired reads in FASTQ format were aligned to the hg19 reference genome using BWA-MEM ^20^ version 0.7.14-r1136. PCR duplicate reads were removed with Picard tools (http://broadinstitute.github.io/picard/) version 1.111 and local realignment of indels was performed using Genome Analysis Toolkit (GATK) ^21^ version 3.8-0 to produce final BAM files (for alignment statistics see File S1 sheet “BAM_files”).

Small variants were defined as “single nucleotide variants” (SNVs) and “small insertions or deletions” (indels) and were called from the final BAM files using GATK HaplotyeCaller ^22^ version 4.1.4.0 in genomic variant call format (gVCF) mode. The resulting gVCF files were jointly genotyped with the GATK commands “GenomicsDBImport” and “GenotypeGVCFs” to produce one multi-sample VCF for the whole sequenced cohort. To calibrate and normalize, we split the cohort VCF by variant type using the “SelectVariants” command from GATK, applied recommended hard filtering to both variant sets using the “VariantFiltration” command, merged the filtered VCFs using “MergeVcfs” command and finally normalized and split multiallelic sites using the “LeftAlignAndTrimVariants” command to produce the final VCF.

SnpEff^23^ and SnpSift^24^ were used to annotate the resulting cohort VCF with variant consequences and information from dbNSFP^25^ version 4.0a. Additionally, we annotated splice prediction scores from SPIDEX/SPANR^26^ version 1.0 and from dbscSNV^27^ version 1.1 and clinical variant assessments from the ClinVar ^28^ database (status 2020-02-10) and from HGMD ^29^ version 2019.3.

The annotated variants were filtered to pass calibration, have an allele frequency < 5% in the cohort and < 1% in gnomAD exomes/ genomes with no homozygotes allowed, not being annotated as (likely) benign, while keeping all variants annotated as (likely) pathogenic in ClinVar. Only variants annotated as high or moderate impact on the gene product or having at least one splicing score predicting aberrant splicing were further analyzed. Compare File S3 sheet “hc-joint”.

Copy number variant (CNV) calling from panel data was performed using CNVkit ^30^ version 0.9.6. The parameters “target-avg-size” was set to 50 and “antitarget-avg-size” to 200.000 to optimize settings for the smaller panel design. The cohort was divided by the sequencing machine (MiSeq vs. HiSeq) and both sub-cohorts were randomly split into two equal sized groups which were used as control cohorts for each other. Resulting per sample CNV calls were annotated with their RefSeq based gene content and aggregated into a cohort list for filtering. Compare File S3 sheet “CNVkit”.

### Variant evaluation and confirmation

Small variants (SNV/indel) were evaluated for their biological plausibility, examined for quality using the IGV browser and classified according to the five-tier variant classification system recommended by the American College of Medical Genetics and Genomics (ACMG) ^31^. For carriers of a (likely) pathogenic variant in *CEP290*, we performed Sanger sequencing to exclude the deep intronic founder variant NM_025114.:c.2991+1655A>G (rs281865192; primers 5’-CATGGGAGTCACAGGGTAGG-3’ and 5’-TGATGTTTAACGTTATCATTTTCCC-3’.

CNVs were visualized with the “scatter” and “heatmap” functions in CNVkit, They were then inspected in the IGV browser to compare their coverage profile with other samples, check the variant allele frequencies (VAF) at variant sites and search for break-point informative split reads. In the sample from individual “Ind_739404” we could identify split-reads supporting the heterozygous *COL4A5* Deletion chrX:g.107731844_107920385del and confirmed the variant with exact breakpoints using allele specific PCR and Sanger sequencing (5’-AATTTGTTGCCTGTCTTTTGC-3’ and 5’-TGCAGAATAAAACCCACACAAC-3’). The deletion (“Ind_958149”) and duplication (“Ind_207310”) affecting the *HNF1B* locus were confirmed using the MLPA kit P241 (MRC Holland, Amsterdam, Netherlands).

### Analysis of the *MUC1-VNTR* region

We analyzed the typical cytosine duplication (“dupC”) located at variable positions in the VNTR between exons 2 and 3 of *MUC1* with an established SNaPshot minisequencing protocol for all archived samples selected for panel sequencing^11^. Additionally, we had designed the panel to include capture probes targeting the *MUC1*-VNTR and included three *MUC1*-dupC positive controls in panel sequencing to enable bioinformatic analysis of this region. We used adVNTR^32^ version 1.3.3 (https://github.com/mehrdadbakhtiari/adVNTR/) with custom settings “frameshift” mode and “vntr_id 25561” to identify indels in this complex genomic region.

### Comparison with published screening data in CKD

To compare our diagnostic yield and exclude potential biases in variant classification, we compared our analysis to the largest currently published sequencing study in CKD^3^. We downloaded all variants from this study directly from ClinVar (SCV000809114 to SCV000809473) as submitted by the authors using a custom R language script. Such downloaded HGVS nomenclature was converted to VCF format using the batch function in VariantValidator (https://variantvalidator.org). We then annotated the resulting VCF file with the pipeline described above for our cohort. Additionally, we annotated whether the respective variant could be detected by our panel using the panel design browser extensible data (BED) file. To harmonize the ACMG classification for our cohort and the Groopman cohort we used the two automated AMCG classification tools integrated in VarSome (https://varsome.com) “ACMG Implementation” and VarSeq v2.2.3 “Sample ACMG Classifier” (Golden Helix, Inc., Bozeman, MT, USA; www.goldenhelix.com) for both variant sets with standard settings. We aggregated multiple (likely) pathogenic variants, as predicted by the ACMG classifiers, per individual in the Groopman cohort and performed 10.000 simulations drawing our final cohort sample size (n=271) from the Groopman cohort. In each simulation we counted how many individuals could be diagnosed by our panel or by exome and how many individuals would have a (likely) pathogenic variant in *COL4A5*. The results of this simulation were then compared with our diagnostic yield using only variants automatically classified as (likely) pathogenic and excluding CNVs and mitochondrial variants. Results were visualized using scatter and violin plots and empirical p-values were calculated by computing how many simulations had a higher or equal yield fraction or COL4 variant fraction, respectively.

### Statistical analyses and plotting

All data regarding cohort, panel content and identified variants were aggregated into Excel (Microsoft Corporation, Redmond, USA) files and are attached as supplementary to this article. These data were imported, analyzed and plotted using R language version 4.1.0 with RStudio IDE version 1.4.1717 (RStudio Inc., Boston, MA, USA). Libraries “broom”, “cowplot”, “DiagrammeR”, “DiagrammeRsvg”, “fs”, “fuzzyjoin”, “ggrepel”, “readxl”, “rsvg”, “tidyverse” and “UpSetR”. Inkscape 1.1 (https://inkscape.org/) was used to adjust Figure 1 and Figure 2 for parts which could not be directly composed in R. Schematic linear gene plots with variant positions represented as lollipops scaled to the variant’s CADD score^33^ were created in R as described previously^34^.

**Figure 2.**
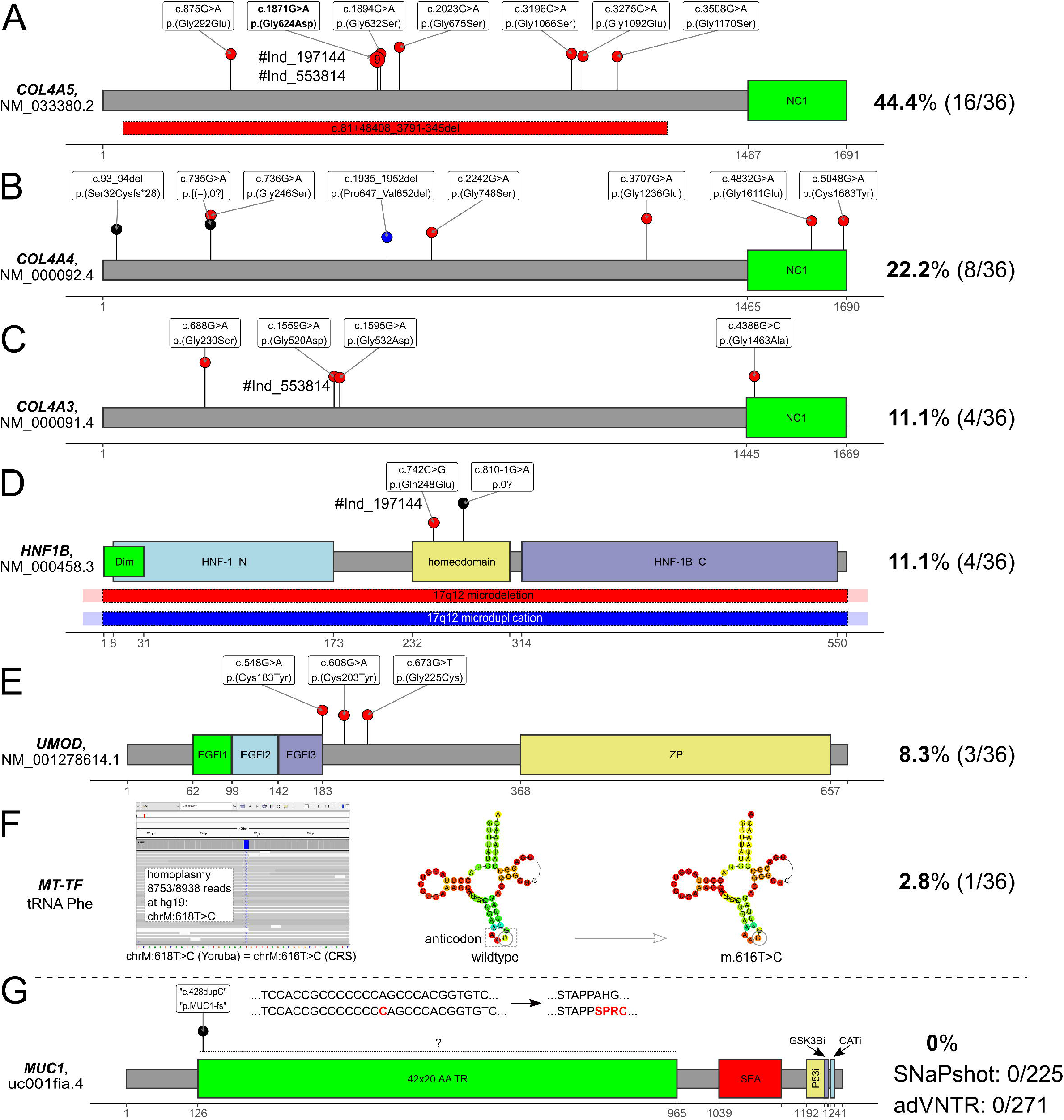
Diagnostic pathogenic variants. Schematic linear protein structure with domains of genes with pathogenic variants identified in the cohort and variant positions marked by lollipops where the length of the segments corresponds to each variant’s CADD score (a computational (“in silico”) metric commonly used to assess the possible pathogenicity of small variants based on an ensemble of annotations like evolutionary conservation). Red dots represent variants, black dots represent variants likely truncating variants, and blue dots represent indels causing in-frame deletions. Red and blue bars with dotted margin represent deletions and duplications, respectively. Individuals with multiple variants identified are linked through the individual pseudonym marked with a “#” under the respective variants. **(A)** In *COL4A5* we identified 15 SNVs and one intragenic deletion. Note that nine unrelated individuals carried the c.1871G>A, p.(Gly624Asp) variant in either hemizygous (six) or heterozygous (three) states. Two individuals (#Ind_197144, #Ind_553814) carried this recurrent missense and another pathogenic variant. **(B)** The eight variants identified in *COL4A4* either affected conserved glycine residues directly through a missense change (four), through an in-frame deletion (one) were likely protein truncating variants (two) or affected a cysteine residue in the C-terminal NC-domain. **(C)** All four variants in *COL4A3* were typical glycine missense changes. One female individual carried the c.1559G>A, p.(Gly520Asp) variant with the recurrent *COL4A5* variant. **(D)** in four individuals we identified variants affecting *HNF1B*. These were a missense variant in the homeodomain, a splice acceptor variant and a genomic deletion and duplication of the 17q12 region, respectively. Deletion breakpoints could not be determined using the sequencing data or MLPA confirmation (red/blue fill overflowing the margin indicating this uncertainty). One female individual carried the c.742C>G, p.(Gln248Glu) variant with the recurrent *COL4A5* variant. **(E)** All three pathogenic variants in *UMOD* were typical cysteine missense variants. **(F)** In the mitochondrial gene *MT-TF*, which encodes the tRNA for phenylalanine, a homoplastic SNV was identified and confirmed. The variant affects the anticodon as predicted through the RNAfold web server ^55^ and has been listed as pathogenic in MITOMAP ^56^. **(G)** Schematic of the MUC1 protein domain structure and the usually unknown position of the typical cytosine duplication (“c.428dupC”) causing a toxic neo-protein in the VNTR region between exons 2 and 3. Bioinformatic search using adVNTR identified no variant and successful “gold standard” SNaPshot in 228 also identified no positive case in the cohort. Grey dashed line used to separate *MUC1* from genes with diagnostic variants in the cohort. Please compare File S2^35^ sheet “domains” for full information on gene protein domains.

The two sided Wilcoxon signed-rank test as implemented in R was used to compare pairwise differences between groups (except for the p-values in the simulation estimated by sampling or when the question could be modelled as a Bernoulli experiment where we used the binomial test).

## RESULTS

### Cohort characteristics

Filtering initially selected 303 patients from the 5,217 individuals of the GCKD cohort (5.8%). 287 (94.7%) DNA samples were of sufficient quality and quantity. Further 16 (5.3%) samples were excluded due to fingerprinting- or sex-mismatch, leaving a final cohort of 271 (89.4%) individuals (Figure 1A). Most individuals fulfilled the inclusion criteria “nephrosclerosis” (94/271 ∼ 34.7%), “IgA nephropathy” (71/271 ∼ 26.2%) or “unknown” etiology (48/271 ∼ 17.7%). 21 individuals (7.7%) were simultaneously in two filtering groups (Figure 1B). The cohort contained 158 males with a median age of 43 years (range 18 - 69 years) and 113 females with a median age of 40 years (range 18 - 66 years), giving a male to female ratio of 1.40 (Figure 1C) which is comparable with the sex ratio in the whole GCKD cohort (3132/2085 ∼ 1.50). Individuals were initially included into the GCKD study following GFR estimation by MDRD study^14^ calculation and CKD-EPI equation based GFR estimates were subsequently performed. We compared these figures for the selected patients, showing little difference between CKD-EPI (median 55.5 ± 24.2 SD mL/min/1.73 m^2^) and MDRD (median 50.0 ± 22.0 SD mL/min/1.73 m^2^) (Figure 1D middle and top panel). The rate of albuminuria at inclusion into the study is expectedly low as we applied search criteria for tubulointerstitial diseases (median 182.1 ± 886.4 SD mg/g creatinine; Figure 1D lower panel). Compare File S1^35^ sheet “cohort” for details per individual.

### High diagnostic yield of 12.5% and genetic spectrum

We identified 36 diagnostic mutations in six genes (Figure 2), which could be classified as type 4 (likely pathogenic) or 5 (pathogenic) variants (Table 1) following the ACMG^31^ recommendations.

**Table 1.**
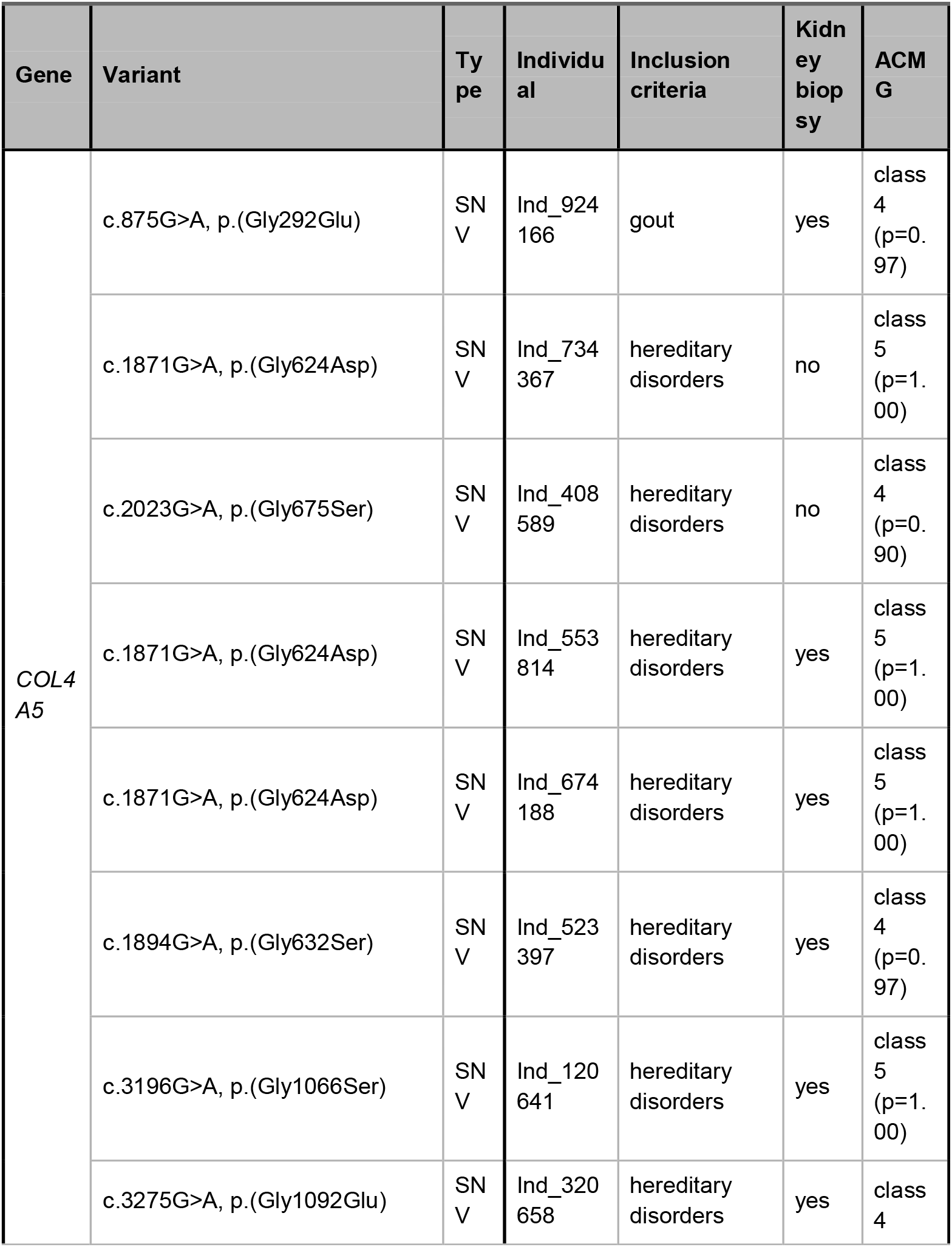
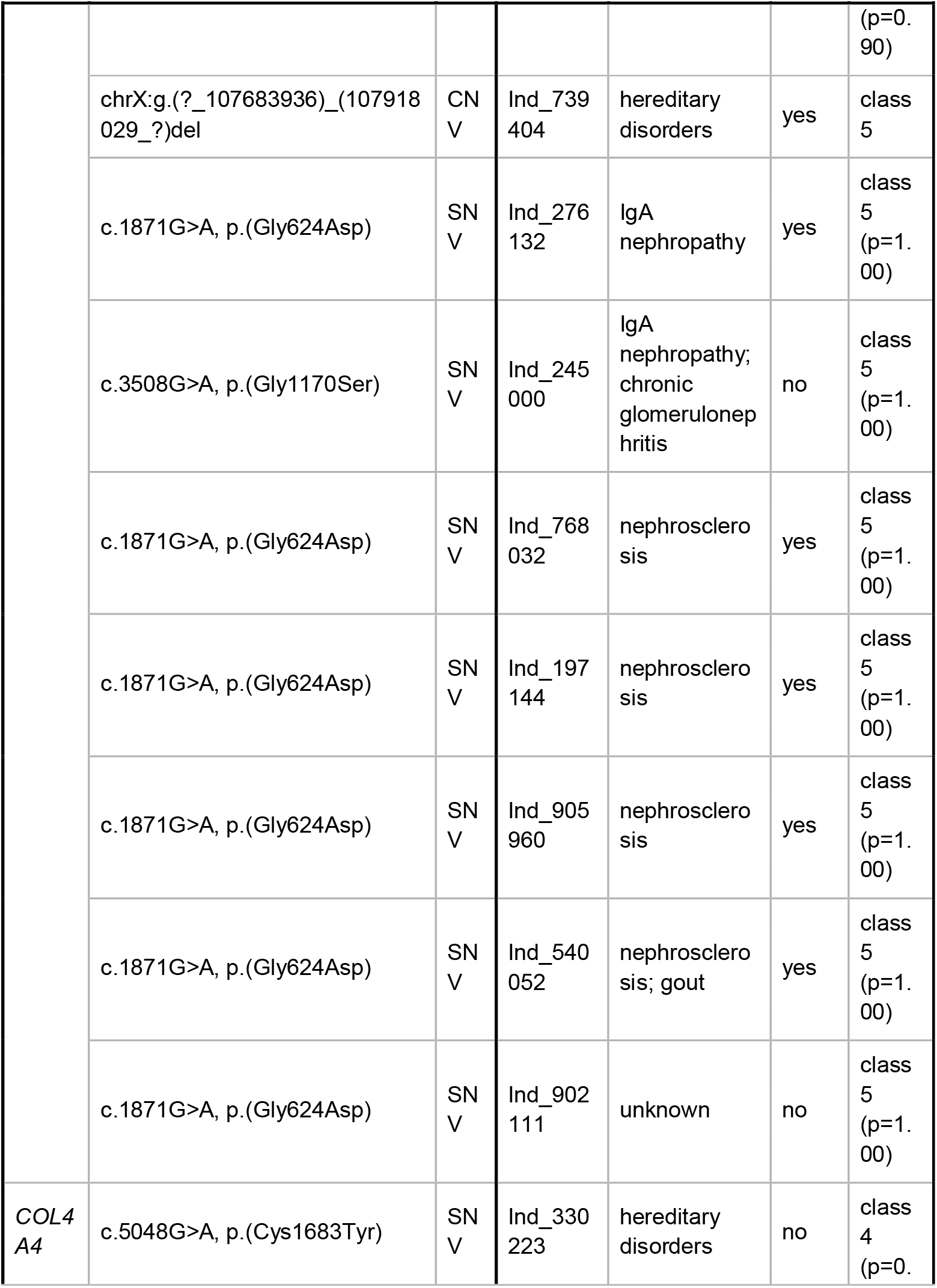
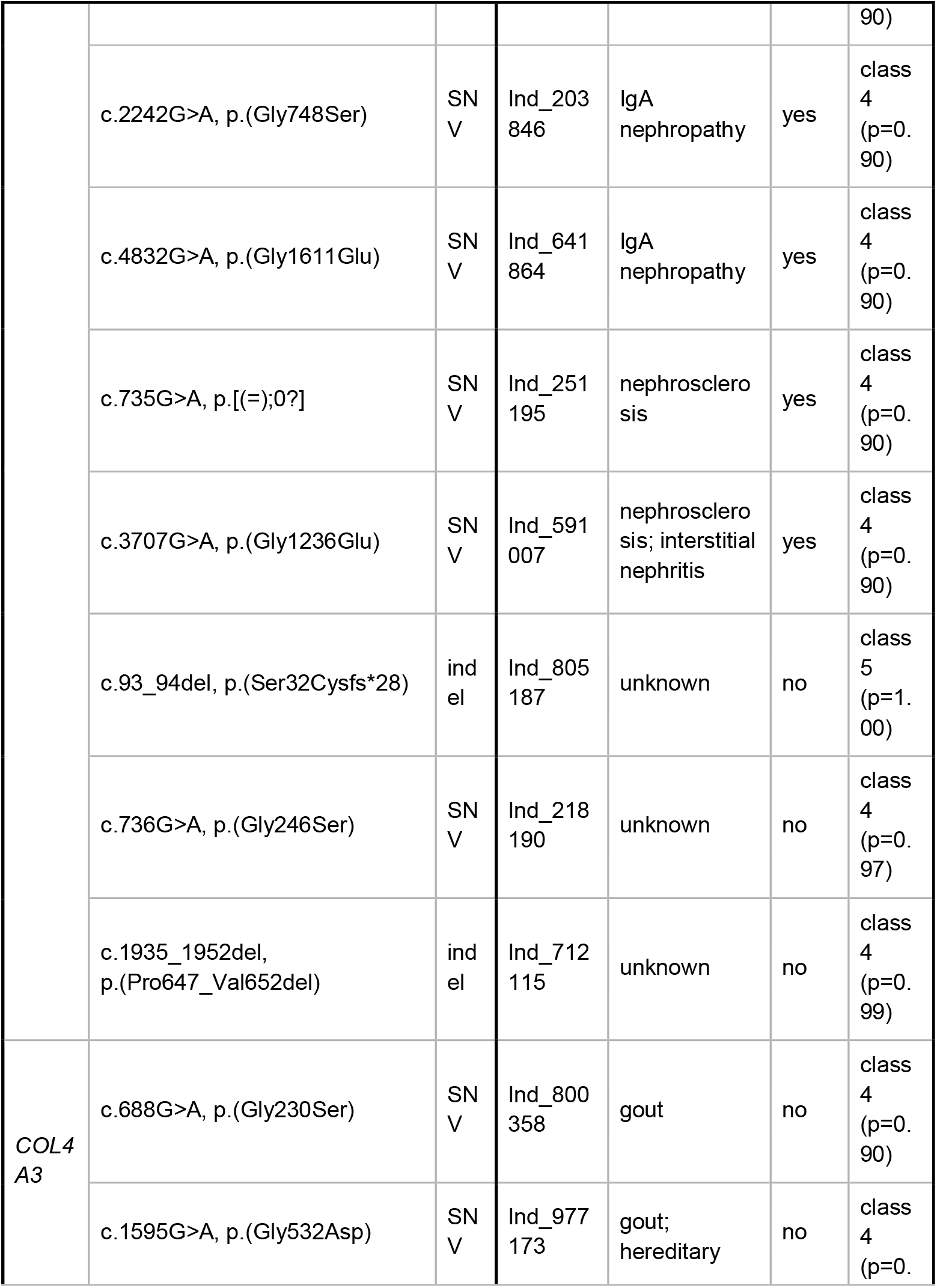
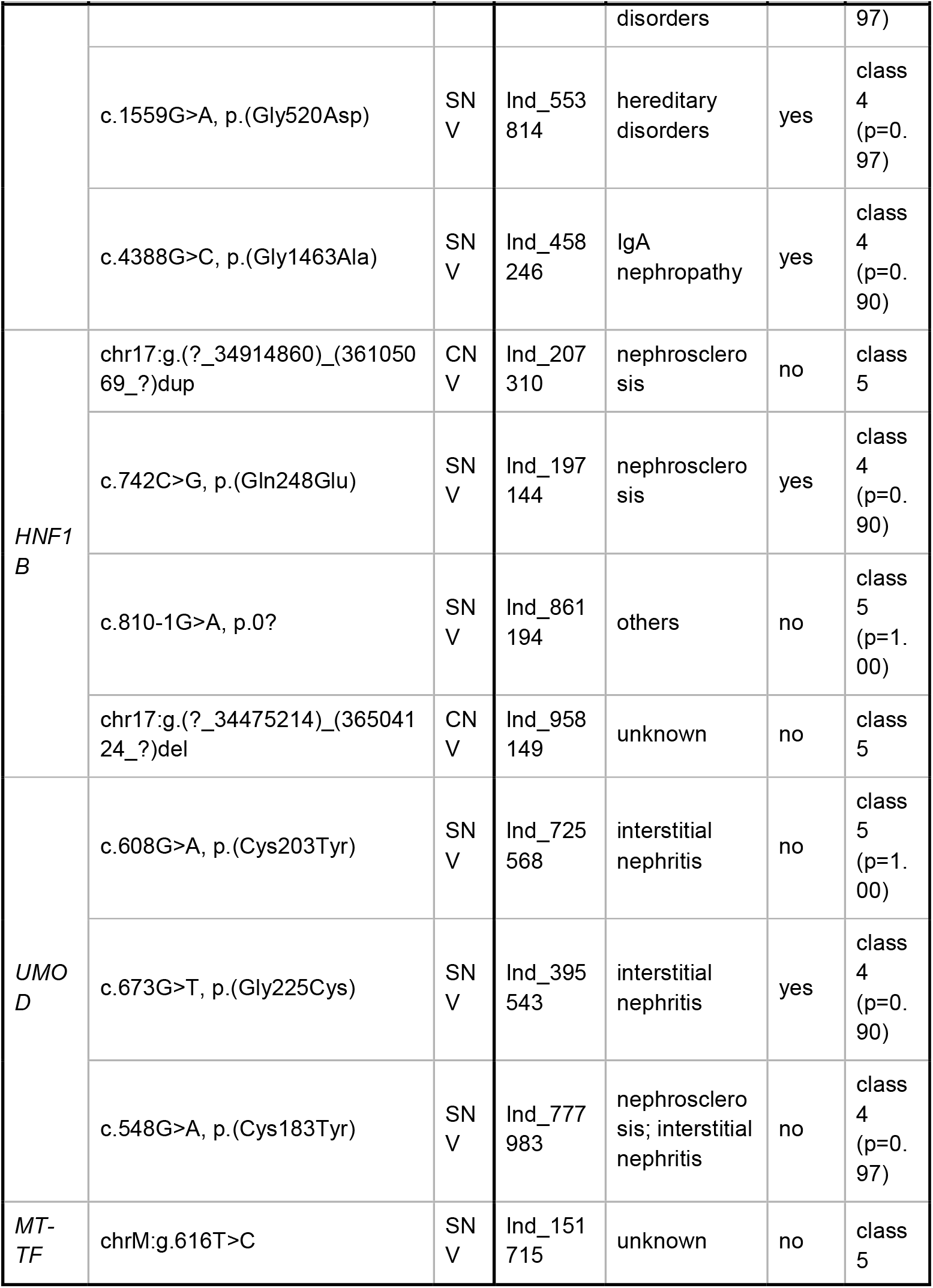
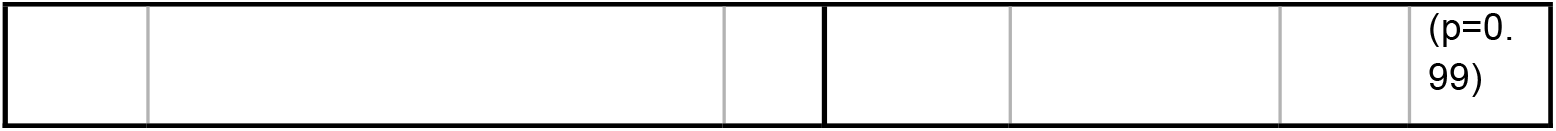
Diagnostic pathogenic variants. List of all individuals who had a (likely) pathogenic variant identified and their diagnostic group/s and whether they had a renal biopsy.

The main focus of our study was to determine the prevalence of ADTKD in a representative cohort of adult patients with CKD. Regarding the classical ADTKD associated genes (*MUC1, UMOD, REN, HNF1B, SEC61A1*), we found three typical cysteine variants in *UMOD* (NM_001278614.1: c.548G>A, p.(Cys183Tyr); c.608G>A, p.(Cys203Tyr); c.673G>T, p.(Gly225Cys)) and a novel missense (NM_000458.3: c.742C>G, p.(Gln248Glu)), and a novel canonical splice variant (NM_000458.3: c.810-1G>A, p.0?) in *HNF1B*. Copy-number analysis additionally identified a duplication and a deletion of *HNF1B*, respectively, which likely represent larger microdeletions/-duplications. No (likely) pathogenic variants were identified in *REN* and *SEC61A1* or in the non-VNTR region of *MUC1*.

In the targeted mitochondrial genome we identified the homoplastic *MTTF* variant m.616T>C, previously described to cause MITKD^36^, in one male individual (“Ind_151715”). An overwhelming number of mutations (28/36 ∼ 77.8%) were identified in the COL4-gene group. Of the 16 diagnostic mutations in *COL4A5*, nine (56.3%) were the previously reported c.1871G>A p.(Gly624Asp) variant (NM_033380.2), which appears to be a relatively frequent founder variant in Europe and is described to lead to a milder course of CKD^37,38^. According to this, the individuals bearing this variant in our study were dispersed throughout Germany and our kinship calculation indicated no recent relatedness. We additionally identified a 188.5 kilobase large heterozygous *COL4A5* deletion in a female individual for which we were able to determine the exact breakpoints from split reads (chrX:g.107731844_107920385del, NM_000495.4:c.81+48408_3791-345del, p.0).

All 36 (likely) pathogenic variants were identified in 34/271 of the analyzed individuals yielding a diagnostic rate of 12.5%. Interestingly, two (2/34 ∼ 5.9%) female patients with the *COL4A5* variant c.1871G>A, p.(Gly624Asp) showed accompanying diagnostic variants in further genes, *COL4A3* (individual “Ind_553814”) and *HNF1B* (individual “Ind_197144”), respectively. Thus, a digenic mode of inheritance or a blended phenotype from two independent disorders can be postulated, which has previously been discussed for a proportion of patients with AS^39^ and is in line with published numbers for multiple diagnostic loci in rare disease patients^40^ and adult CKD patients^3^. Reported variants with the respective patient’s clinical criteria are listed in Table 1. Table 2 shows additional variants of unknown significance identified and Table 3 lists the (likely) pathogenic variants identified in nephronophthisis genes.

**Table 2.**
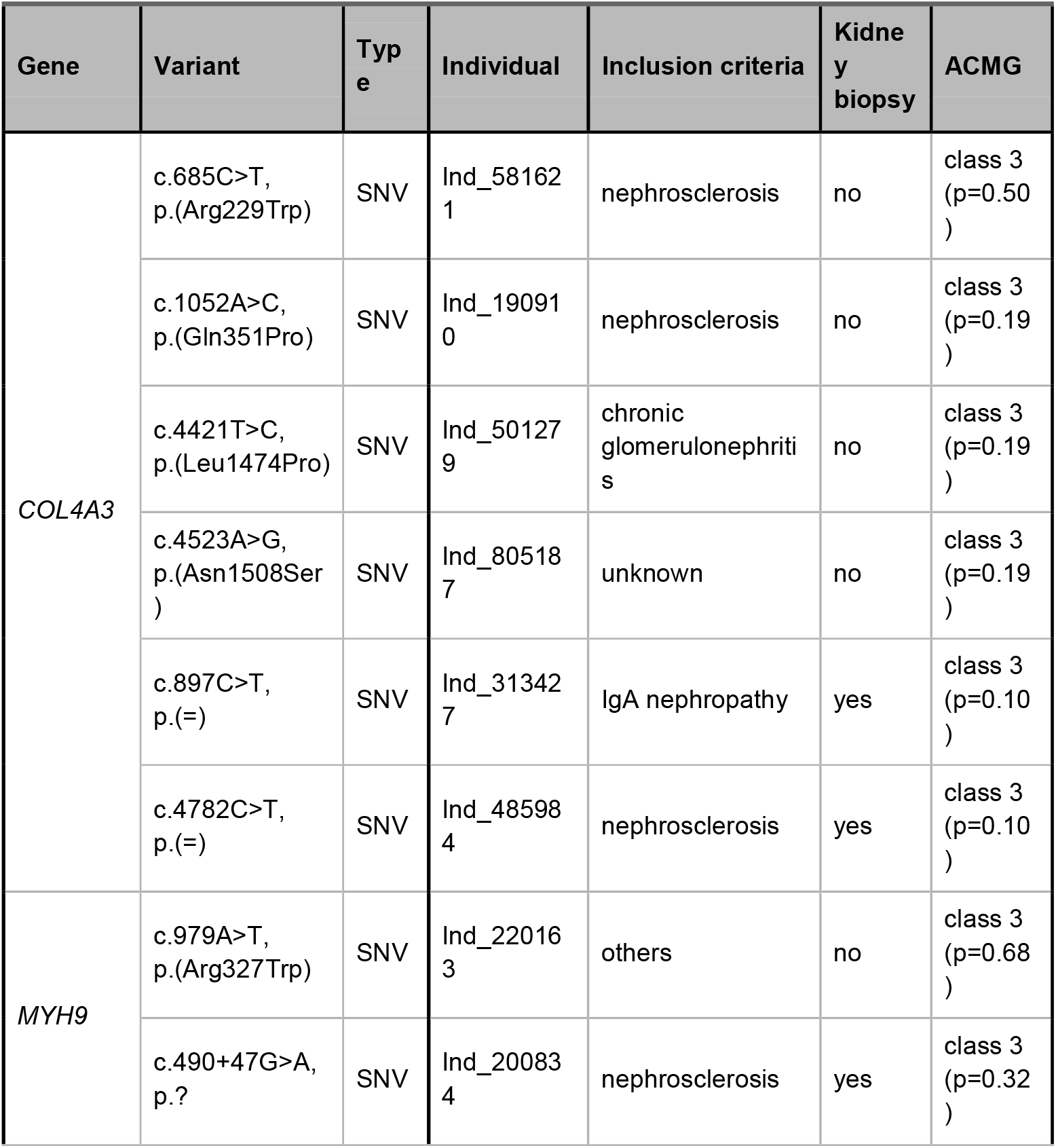
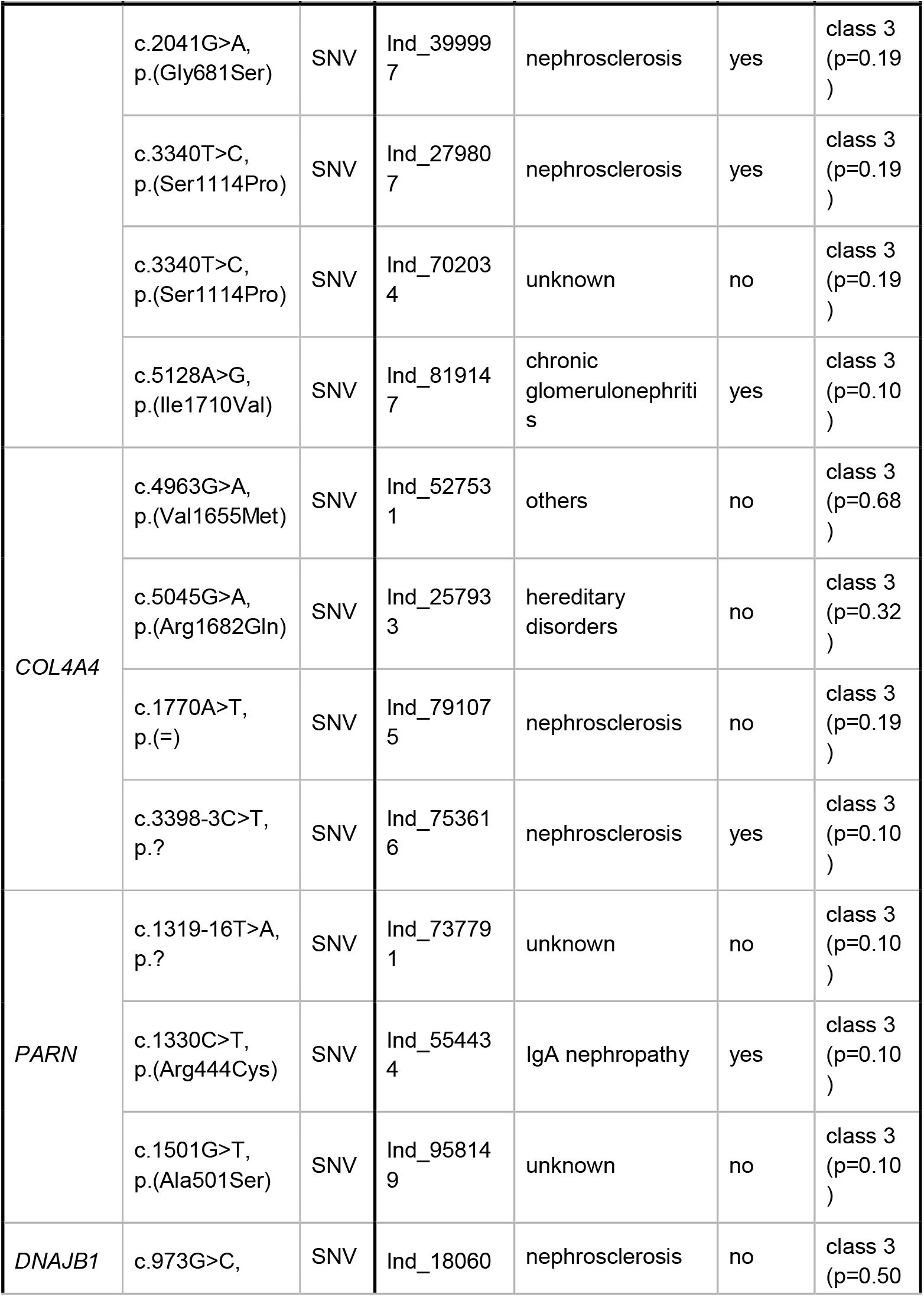
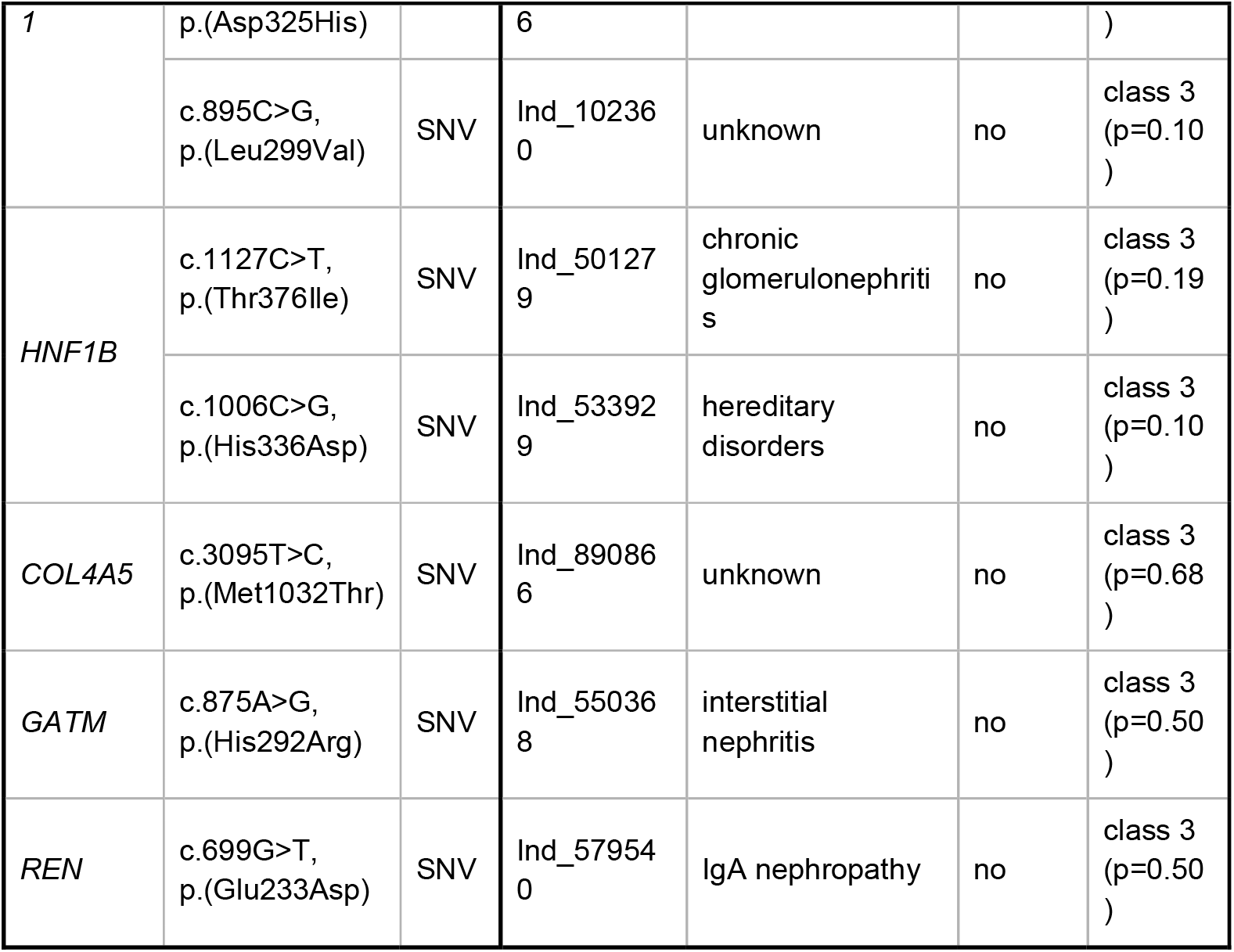
Additional variants of unknown significance. List of all individuals who had a VUS identified and their diagnostic group/s and whether they had a renal biopsy.

**Table 3.**
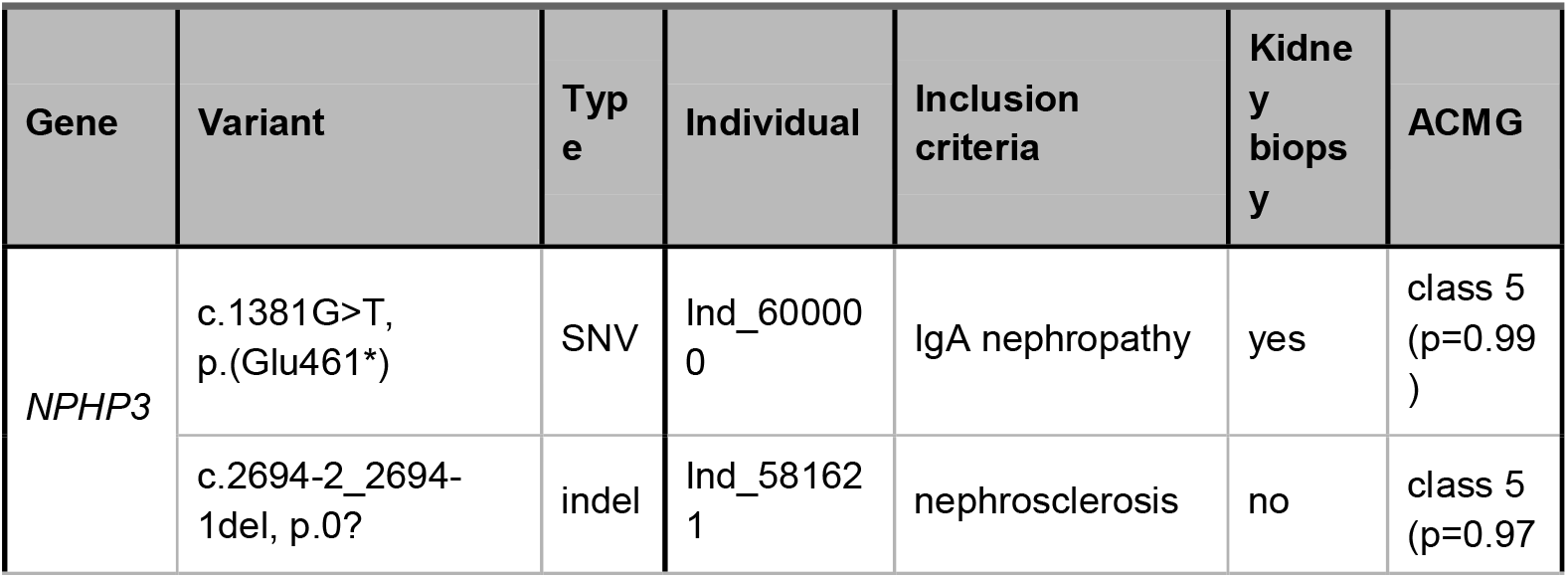
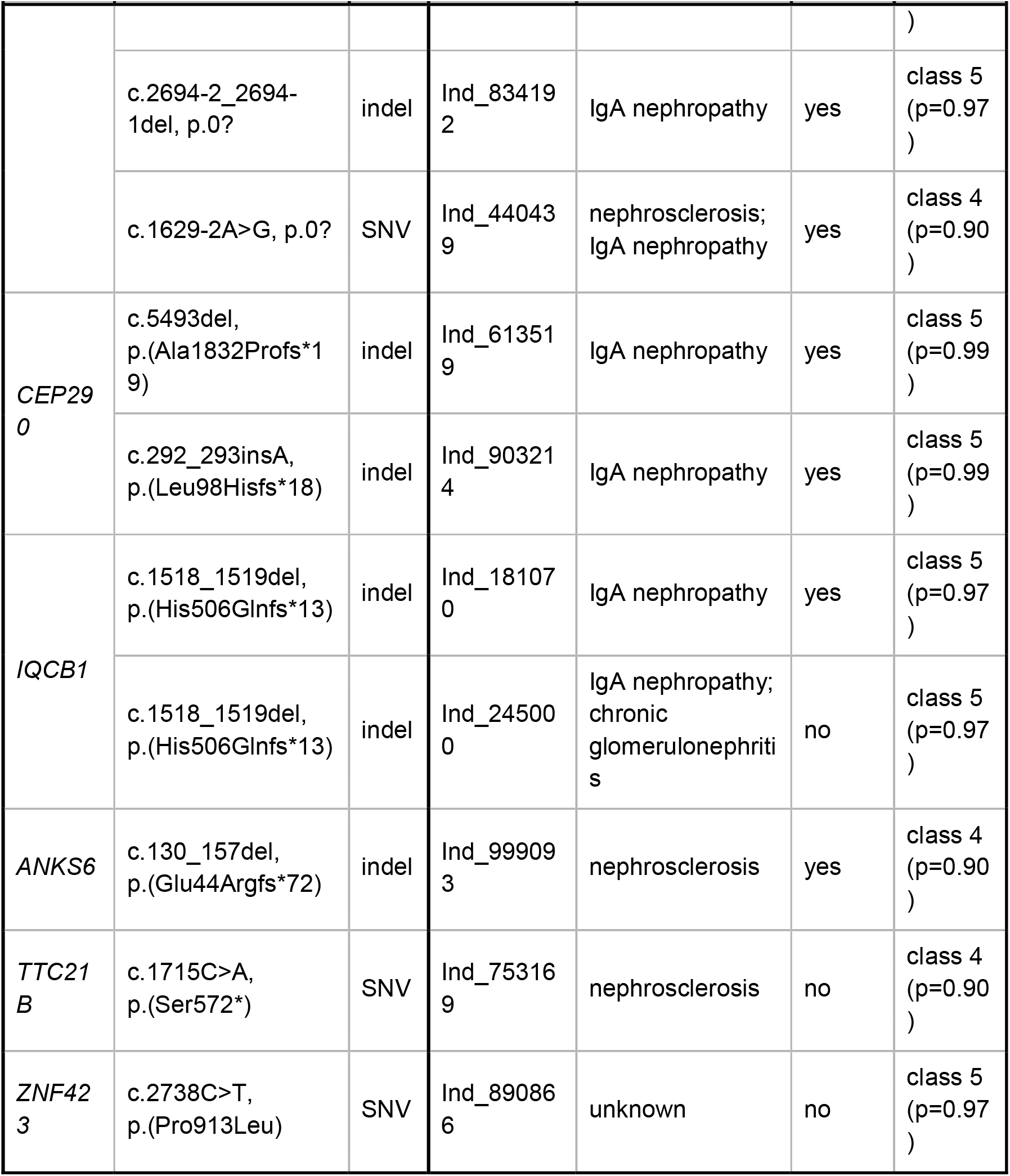
Nephronophthisis carrier variants. List of all individuals in which a heterozygous (likely) pathogenic variant in 17 Nephronophthisis genes was identified together with their diagnostic group/s and whether they had a renal biopsy.

### Relation of clinical criteria and genetic diagnosis

Having identified the patients with an underlying genetic disease, we re-analyzed and correlated the clinical information that was available in the GCKD database. As could be expected, the group “hereditary disorders” harboured the highest rate of patients with diagnostic mutations (10/17 ∼ 58.8%), followed by the groups “gout” (4/9 ∼ 44.4%) and “interstitial nephritis” (4/23 ∼ 17.4%) (Figure 3A). The large groups of “nephrosclerosis” and “IgA nephropathy” display lower rates of diagnostic hits with 8.4% (8/94) and 7.0% (5/71), respectively. Assuming an equal diagnostic rate for all categories as null hypothesis, only the categories “hereditary disorders” (p ∼ 0.000014; binomial test) and “gout” (p ∼ 0.023; binomial test) showed significant enrichment for genetic findings. The group “hereditary disorders” would remain significant when correcting for multiple testing at a threshold of 0.005/9 (∼ 0.0056). By far the most diagnostic mutations involve one of the three COL4-genes, which are the sole mutations in the groups “gout”, “hereditary disorders” and “IgA nephropathy” (Figure 3B). The diagnostic groups “interstitial nephritis”, “nephrosclerosis” and “unknown” show a higher rate of genetic heterogeneity. However, the numbers are too small to speculate about a systematic effect.

**Figure 3.**
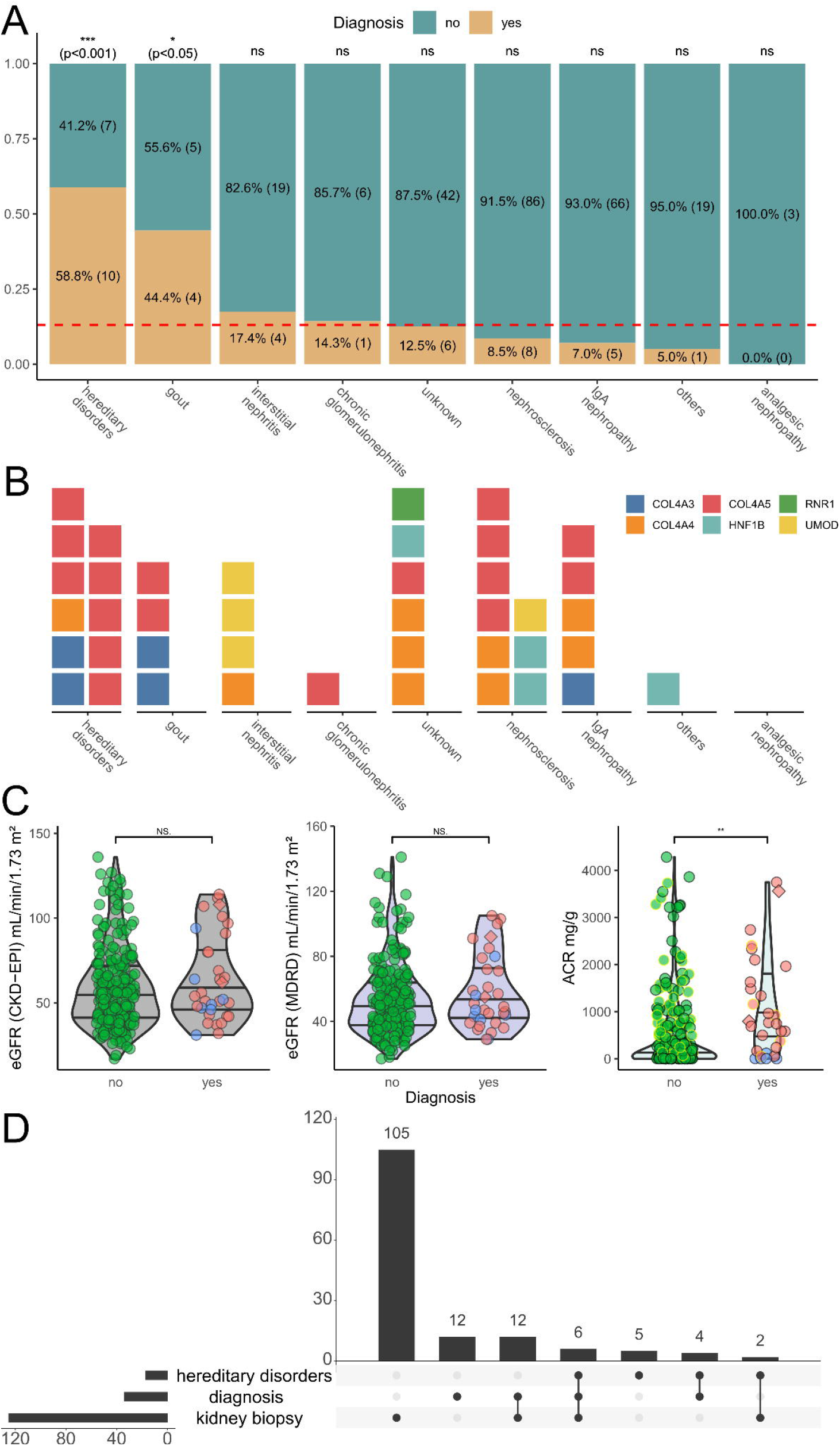
Pathogenic variants by clinical criteria. **(A)** Stacked bar plot indicating the fraction of individuals with a pathogenic variant identified split by the nine clinical filtering criteria. As some individuals fulfilled multiple criteria we split them by diagnosis. This resulted in 292 total combinations of individuals and clinical criteria and 39 such combinations for individuals with a genetic diagnosis. To test whether certain criteria are enriched for genetic findings, we calculated p-values assuming an equal diagnostic rate of 39/292 in a simple Bernoulli experiment using a binomial test. Categories “hereditary disorders” (p ∼ 0.000014) and “gout” (p ∼ 0.023) showed nominally significant enrichment. The “hereditary disorders” category remained significant after adjusting for multiple testing. **(B)** Waffle plot comparing the nine filtering criteria and the gene in which a variant has been identified. Note that the 39 combinations of individuals and criteria are now also split by gene, because two individuals in the cohort had multiple pathogenic variants, resulting in 41 combinations. The two significant categories from A are all explained through variants in the COL4-genes. Interestingly all three *UMOD* variants identified fall in the “interstitial nephritis” category, with one of them additionally classified as “nephrosclerosis”. Variants affecting *HNF1B* are either dispersed through four categories with none of them in the hereditary category, confirming both the variability in the *HNF1B*-associated disorders and the often sporadic nature of the CNVs (17q12 microdeletion/ -duplication syndromes). Compare File S3^35^ for full variant details. **(C)** Violin and scatter plots comparing the kidney function parameters from Figure 1D between individuals with a genetic variant identified (34) or not (237; green circles). Individuals with a COL4-variant are presented in red and with variants in other genes in blue. Individuals with two variants are marked as diamonds. Individuals with IgA nephropathy are marked with yellow margin (compare also Figure S2). The ACR at GCKD study inclusion is significantly higher in individuals with a genetic variant identified (two sided Wilcoxon signed-rank test). **(D)** Upset plot showing the overlaps for individuals with a suspected “hereditary diagnosis” (as used for filtering), our finding of a pathogenic variant (“diagnosis”) and kidney biopsies performed. Overall only in six individuals with a confirmed diagnosis a kidney biopsy had been performed previously which likely raised the suspicion of an underlying genetic disorder. In 12 individuals with kidney biopsy where we identified pathogenic variants no suspicion of a hereditary disease was issued.

Comparison of the GFR at inclusion into the GCKD study between the group of patients where a genetic diagnosis was identified with the rest of patients did not show a difference (Figure 3C, left and middle panel). In contrast, the albuminuria at inclusion into the study was significantly higher in the genetically determined group, which is an effect exclusively caused by the patients with AS (Figure 3C, right hand panel).

Next, we were interested in the contribution of previous kidney biopsies for the clinical evaluation, since the kidney histology is not informative for the diagnosis of ADTKD ^6,7^, but in contrast could be helpful in recognition of COL4-diseases. Figure 3D shows that a biopsy was taken in 46.1% (125/271) of the selected patients before inclusion into the GCKD study. Interestingly, this rate was similar with 47.1% (8/17) in patients that were classed into the group of “hereditary disorders” and with 52.9% (18/34) in the group of patients with a diagnostic mutation. Considering only the patients with an identified COL4-disease, 63.0% (17/27) of this group were biopsied, but only three patients (11.1%) were previously marked with a suspected diagnosis of AS in the GCKD files (two biopsied, one not biopsied). All these three patients were correctly positioned by the nephrologists into the group “hereditary disease”, where the rest of patients in this disease group were commented as “unspecified”. Therefore, for the patients analysed in our study the kidney biopsy does not appear to have been of any direct diagnostic value, unless for exclusion of another disease.

### Comparison with published CKD cohorts confirms high diagnostic rate

Compared to previous studies of adult CKD cohorts, our diagnostic yield of 12.5% (34/271) is relatively high and comparable to exome sequencing, despite the relatively small number of genes in our design and exclusion of *PKD1*/*2* associated disease. To test for the generalizability of this observation, we compared our diagnostic yield to the currently largest exome sequencing study in adults with CKD by Groopman and colleagues^3^. As this study did not analyze CNVs and mitochondrial variants, we also only included small variants in the autosomes and gonosomes from our study (30/271 ∼ 11.1%; excluding CNVs and mitochondrial variants) for the comparison. After harmonizing both our and the AURORA and CUMC cohorts^3^ using the same annotations, we performed a simulation where we randomly selected 271 individuals from the 3,315 individuals reported with diagnostic variants by Groopman et al. and then counted whether the respective variant reported would be detectable by our analysis. The simulation indicated that our diagnostic yield of 11.1% is very unlikely by chance (estimated p-value < 0.0001) (Figure 4A), and this indicates enrichment through our filtering (compare Figure 4B).

**Figure 4.**
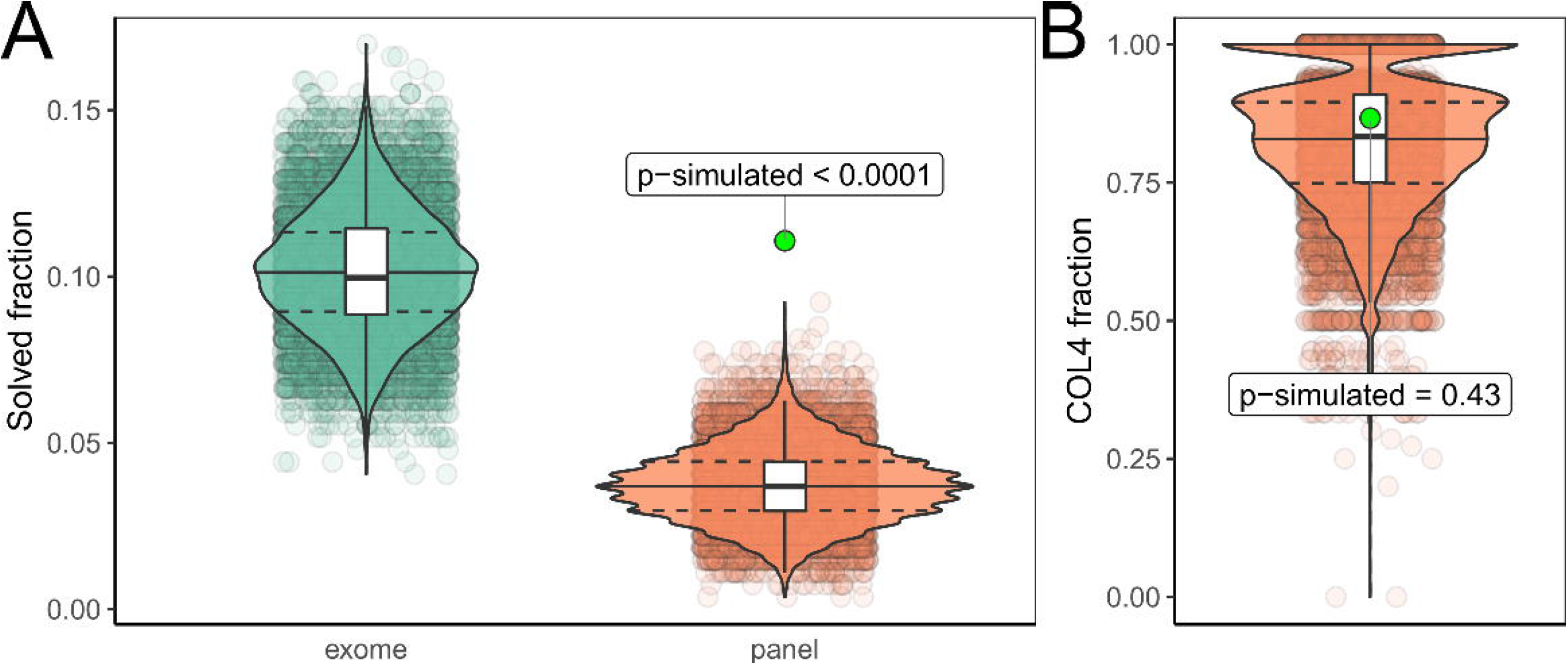
Diagnostic yield in panel and exome. **(A)** Violin and scatter plots of 10.000 simulations randomly drawing 271 individuals from the 3,315 individuals reported by Groopman et al. and subset whether the reported variants would be detectable by exome (green) or our targeted panel (red). Estimated p-value for the yield in our cohort (green dot) < 0.0001. To exclude differences in variant classification between the two studies, we classified both our and all variants from the Groopman study using two automated ACMG classifiers which excluded all variants not classified as (likely) pathogenic from both cohorts. This gave similar results to the first simulation and thus excluded systematic differences in manual variant classification causing our higher yield (compare Figure S1). **(B)** To exclude unexpected enrichment for Collagen-4 genes, we further compared the fraction of COL4-gene variants in these simulations which showed no significant difference (p-simulated ∼ 0.43) to the fraction (26/30 ∼ 86.7%) observed in our cohort. Therefore, one could consider the COL4-variants as background, which would leave five small variants in ADTKD genes in our cohort, representing an enrichment of ∼ 5.1 fold when compared to the Groopman cohort (calculation: (5/271) / (12/3,315)). Compare Figure S1 for automated classification results. Compare File S4^35^ for full simulation results.

## DISCUSSION

CKD is frequent disease, affecting more than 10% of the global population, that is strongly associated with adverse prognosis and has a profound socioeconomic impact.^41,42^ In the last decade genetic diagnostics have greatly improved, which has led to the recognition of a relevant burden of hereditary causes amongst patients with CKD. In parallel, international initiatives promote targeted treatment developments for rare diseases. The aim for “precision medicine” therefore thrives for an accurate diagnosis and a effective targeted therapy.^1,2,43^

ES on a clinical basis in every patient with CKD is not (yet) realistic, is not standardized (e.g. different commercial designs and bioinformatic pipelines) and has diagnostic gaps for several kidney disorders. Thus, algorithms need to be defined to decide which patient should be offered genetic testing and which combinations of ES and specialized targeted analyses will result in highest diagnostic yields while being as economical as possible for the healthcare system. Possible criteria to undertake genetic analysis would be young CKD onset, disease type and a positive family history as well as the existence of extrarenal, syndromic features.^4^ To date, the largest genetic study published on CKD patients analyzed a virtual panel of 625 genes associated with kidney disease on an exome platform^3^. In this study 63% of diagnostic mutations were restricted to six genes (*PKD1*/*2, COL4A3*/*4*/*5, UMOD*). Therefore, on a clinical basis it appears appropriate to restrict the number of analyzed genes. We here used a panel of merely 29 genes to investigate the prevalence of hereditary tubulointerstitial diseases. Our rate of diagnostic findings was higher compared to Groopman et al. (12.5% vs. 9.3% or 10.1 when including their “putatively diagnostic variants”)^3^, which we interpret as confirmation of successful filtering criteria (Figure 4).

The majority of our diagnostic findings were amongst the COL4-genes (Figure 2), which would not normally account for tubulointerstitial but glomerular diseases. However, COL4-diseases have been extensively reported as frequent unexpected diagnoses in patients with focal segmental glomerulosclerosis upon renal histology^44^ or broad population based analyses,^3,45^ where previous erroneous diagnoses may have taken place. Therefore, we decided to include the COL4-genes. The rate of COL4-mutations may be lower in other populations since about one third of the pathogenic COL4-variants we found were the *COL4A5* hotspot mutation c.1871G>A (p.Gly624Asp) (10/28 ∼ 35.7% here vs. in the Groopman study 9/108 ∼ 8.3%), with a high frequency in central European populations^37,38^. Interestingly, looking back into the original entries of the GCKD database, of the 28 patients with a diagnostic COL4-variant, only three were previously suspected to have AS. Thus, the majority of almost 90% of COL4-diseases were clinically not recognized. Analysis of the defined AS patients for proteinuria showed a significant difference for a moderate proteinuria as compared to the group without a genetic diagnosis (Figure 3C). Therefore, recognition of proteinuria could sensitize nephrologists towards COL4-diseases and encourage a restricted diagnostic workup. Overall, our analysis confirmed previous studies showing a high background rate of COL4-variants in CKD cohorts. Considering that two individuals with a pathogenic COL4-variant had a second pathogenic variant (2/34 ∼ 5.9%) and thus a dual diagnosis, it seems sensible to perform a broader search in patients with a COL4-variant, especially if the affected person shows an atypical disease course or additional features.

Further rather difficult diagnostic groups, prone to faulty classification could be “nephrosclerosis” and “IgA nephropathy”, where our analysis yielded diagnostic mutations in 8.5% and 7.0% of patients, respectively (Figure 2A). Although these groups showed no statistically significant enrichment, when compared to the baseline diagnostic yield in our cohort, they could motivate clinicians to look more careful at individual patients before diagnostic classification. Naturally, the great majority of CKD patients will show arterial hypertension and often it will not be clear if this is the cause or sequel of CKD. Similar challenges can be met with the histological diagnosis of “IgA nephropathy”, which can be found in a substantial fraction of the (healthy) population.^46–48^ Therefore, parallel and possibly more severe diagnoses such as ADTKD can be overlooked.^49^

Our study investigated a diagnostically particularly difficult group of patients with hereditary tubulointerstitial diseases. Since patients suffering from ADTKD usually reach ESRD between the 3rd and 6th decade of life ^6,7^ and the GCKD inclusion criteria was CKD stage 3 ^14^, we set the age cut-off for most leading diagnoses to 40 or 50 years of age (see Methods). This stringent age-related cut-off accepts that single patients may be missed with an exceptionally mild phenotype, which however is rarely the case with ADTKD.^50,51^ The comprehensive diagnostic difficulties are clinical and histological but also methodological in terms of molecular genetics. As such, respective candidate genes may have frequent CNVs (i.e. *HNF1B*), are not contained in usual genetic screens (mitochondrial genome) or show complex repeat structures (i.e. *MUC1*). In the absence of a family history it is very difficult to raise a clinical suspicion of these diseases. Therefore, we suspect that patients with sporadic disease will hardly be recognized. Thus, the prevalence of these diseases are not known to date. We identified seven mutations in known genes for ADTKD which represent 2,6% (7/271) of the sequenced cohort. Interpolating to the total GCKD cohort, while assuming complete enrichment through our criteria, this would mean a prevalence of 0.13% (7/5,217). Interestingly, this figure is similar to another recent study with the estimate of 0.54% for patients with ADTKD in the complete ESRD cohort of Ireland ^52^ and the diagnostic yield for ADTKD variants reported by Groopman et al. with 0.39% (13/3,315). Importantly, none of the 7 individuals with diagnostic ADTKD mutations were originally grouped as suffering from a “hereditary disorder”. These patients were originally classed as “nephrosclerosis” (3x), “interstitial nephritis” (3x), “others” (1x) and “unknown” (1x) (see Table 1). Therefore, we presume that the majority of these diseases could be sporadic.

While we present a detailed analysis of the prevalence of hereditary tubulointerstitial kidney diseases, it is important to consider confounders. First, any selection strategy has the potential to miss patients. Thus, the true figure of hereditary disease will presumably be higher than our results. Second, we performed a screening limited by a customized gene panel which can miss other causative mutations. However, the focus of our study was ADTKD and related diseases, which were tested exhaustively. Third, we classed the variants following the recommendation of the ACMG^31^, where class 3 VUS are not contained in our yield calculations. We performed a detailed analysis of these VUS (Table 2). However, it currently remains unknown how many of them in fact are the reason for CKD in individual patients and further population and functional studies (e.g. saturation mutagenesis) will be needed to elucidate their effects. Fourth, we did not include the genes recommended to be reported as secondary findings^53^, which are expected in ∼ 1% of the population and are of clinical relevance especially for CKD patients with chronic dialysis or immunosuppression^4^. In summary, ADTKD/MITKD are quite rare in the CKD population. With limitations in financial resources, it is probably not justified to broadly perform targeted ADTKD diagnostics in the clinical routine. This is particularly true for ADTKD-*MUC1*, where testing for the “dupC”-mutation using SNaPshot is laborious and did not lead to a single hit here. On the other hand, our bioinformatic assessment of the targeted VNTR region showed complete agreement. Also, when clinical criteria are present and a clear autosomal dominant pedigree evident, the rate of diagnostic mutations for ADTKD is reasonably high.^50,54^ Based on these considerations, our and other’s results and experience from rare disease studies we recommend a clinically enhanced ES design paired with customized bioinformatics (Figure 5A) and an iteration of genetic diagnostics and research re-evaluation (Figure 5B). Only by establishing such comprehensive workflows in centers for rare kidney diseases will we be able to improve diagnostics, gather further knowledge on each genetic CKD entity and finally improve outcome.

**Figure 5.**
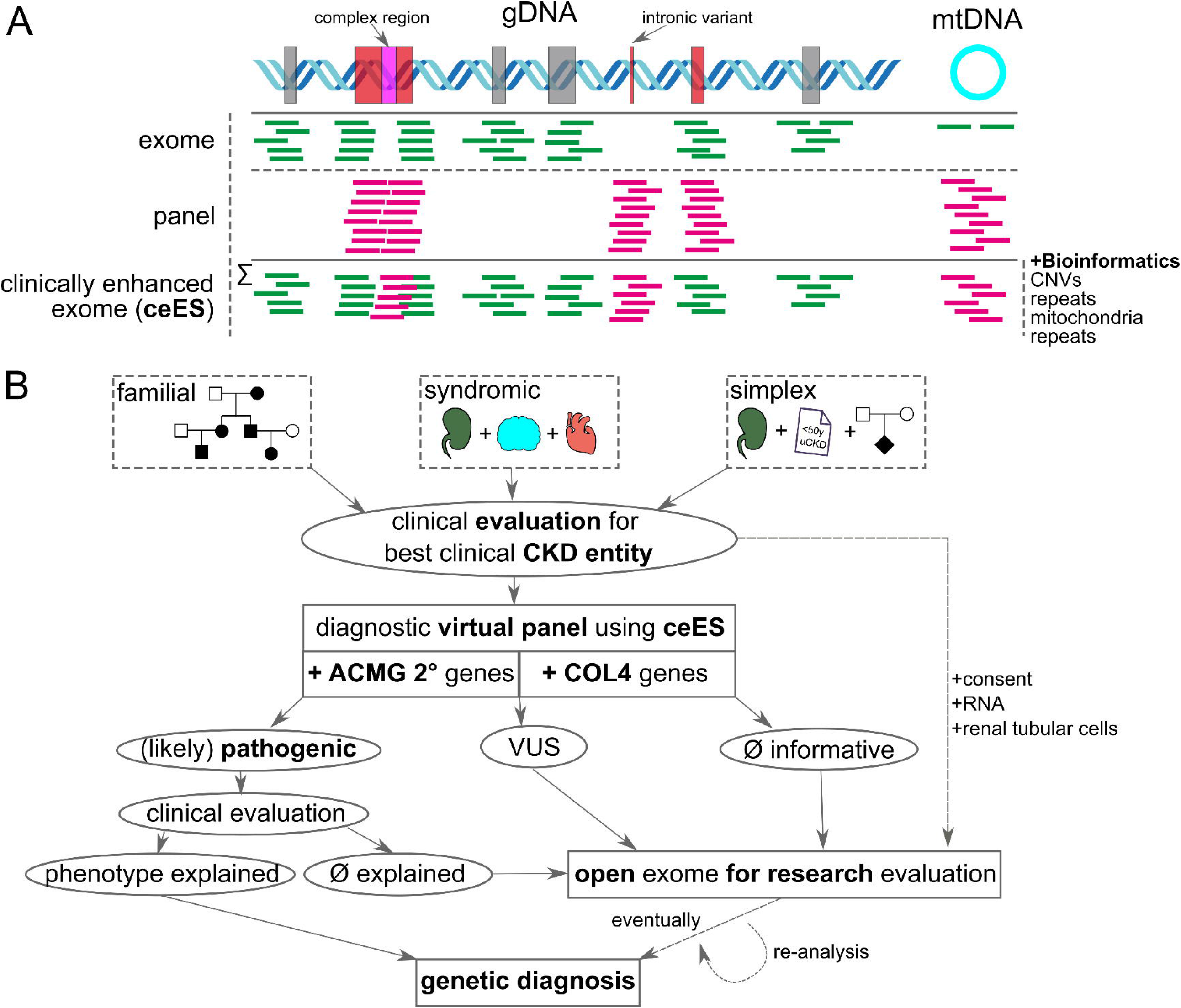
Proposed clinically enhanced exome design and evaluation workflow. (**A**) Schematic figure explaining short read sequencing based panel and exome sequencing (ES) and their respective advantages. ES covers all coding exons but has gaps in complex regions (like the *MUC1*-VNTR or *PKD1* duplicated exons), may miss clinically relevant intronic variants and has low coverage for mtDNA. Custom panels in contrast can be designed to have high coverage of these regions but would need to be iteratively redesigned and re-sequenced for each possible disease entity. An ES target design enhanced through expert knowledge (several companies nowadays offer adding custom capture probes) allows adapting the design to the respective diagnostic needs (ceES). **(B)** Our proposed workflow to select patients for genetic diagnosis is based on positive family history, syndromic disease (e.g. multiple organ systems affected) and isolated simplex cases without secondary cause of CKD younger than 50 years. Genetic diagnostics should be based on clinically selected virtual panels and include ACMG recommended secondary findings and COL4-genes. Depending on the outcome and whether eventual variants explain the phenotype of the patient the ceES data should be opened to research analysis enhanced by possible RNA analyses and functional tests using e.g. renal tubular cells to finally reach a genetic diagnosis.

## Data Availability

All data is available through Zenodo.

https://doi.org/10.5281/zenodo.5516388

## AUTHORS CONTRIBUTIONS

M.W., B.P. and A.B.E. conceived the initial study concept. M.W. curated the clinical criteria and filtered/ selected the cohort. A.B.E., M.W., and B.P. created the panel gene list. A.B.E., M.S., and K.U.E. collected the samples from the GCKD cohort and provided clinical and chromosomal microarray data. A.B.E. coordinated panel design, DNA sample processing and all sequencing. S.U. and B.P. performed tertiary bioinformatic analyses. J.P. and V.B. developed adVNTR and analyzed the *MUC1*-VNTR panel data. K.S., K.K. and A.B.E. performed the *MUC1*-SNaPshot analysis. U.A. aided in the validation and classification of mitochondrial variants. A.R. and C.K. performed variant validation and aided in quality control. B.P. performed variant calling, annotation, classification and upload to public databases. B.P. analyzed the variant and clinical data and created all figures/ tables and the Supplementary materials. B.P. and M.W. wrote and edited the manuscript. All authors reviewed and commented on the final draft manuscript.

## ACKNOWLEDGEMENTS

We thank all involved individuals for participating in the GCKD study. We also thank the large number of nephrologists for their support of the GCKD study (list of nephrologists currently collaborating with the GCKD study is available at http://www.gckd.org). The GCKD Investigators are listed in the Supplementary Notes. We would like to thank Susanne Becker for valuable assistance with data analysis of GCKD. We would like to thank Petra Rothe, Angelika Diem and Daniela Schweitzer for excellent technical assistance. We thank Uwe Ahting (Institute of Human Genetics, Technische Universität München, Munich, Germany) for aiding with the interpretation of mitochondrial variants.

## SUPPLEMENTAL MATERIAL

### Supplementary Notes

Contains extended acknowledgements, supplementary methods and results, two supplementary figures, web resource links and abbreviations used.

**File S1**

Cohort characteristics, sequencing quality parameters and fingerprinting results. File is available through Zenodo https://doi.org/10.5281/zenodo.5516388.

**File S2**

Sequencing panel design/ content with information on gene domains used for Figure 2. File is available through Zenodo https://doi.org/10.5281/zenodo.5516388.

**File S3**

Information on small variants, CNVs and *MUC1* analyses (SNaPshot and adVNTR). File is available through Zenodo https://doi.org/10.5281/zenodo.5516388.

**File S4**

Curated variant and individual data from the Groopman study with results of the simulation for Figure 4. File is available through Zenodo https://doi.org/10.5281/zenodo.5516388.

