## Supplementary notes for "Prevalence of hereditary tubulointerstitial kidney diseases in the German Chronic Kidney Disease study"

**Running headline:** Prevalence of ADTKD in the GCKD study

**CORRESPONDING AUTHOR:**
Prof. Dr. Michael S. Wiesener, M.D.
Department of Nephrology and Hypertension
University Hospital Erlangen
Friedrich-Alexander-University Erlangen-Nürnberg
Ulmenweg 18
91054 Erlangen, Germany


**Extended acknowledgements**

Current GCKD investigators and collaborators with the GCKD study are: University of Erlangen-Nürnberg: Kai-Uwe Eckardt, Heike Meiselbach, Markus P. Schneider, Mario Schiffer, Hans-Ulrich Prokosch, Barbara Bärthlein, Andreas Beck, André Reis, Arif B. Ekici, Susanne Becker, Ulrike Alberth-Schmidt, Anke Weigel; University of Freiburg: Gerd Walz, Anna Köttgen, Ulla T. Schultheiß, Fruzsina Kotsis, Simone Meder, Erna Mitsch, Ursula Reinhard; RWTH Aachen University: Jürgen Floege, Turgay Saritas; Charité, University Medicine Berlin: Elke Schaeffner, Seema Baid-Agrawal, Kerstin Theisen; Hannover Medical School: Hermann Haller; University of Heidelberg: Martin Zeier, Claudia Sommerer, Johanna Theilinger; University of Jena: Gunter Wolf, Martin Busch, Rainer Paul; Ludwig-Maximilians University of München: Thomas Sitter; University of Würzburg: Christoph Wanner, Vera Krane, Antje Börner-Klein, Britta Bauer; Medical University of Innsbruck, Division of Genetic Epidemiology: Florian Kronenberg, Julia Raschenberger, Barbara Kollerits, Lukas Forer, Sebastian Schönherr, Hansi Weissensteiner; University of Regensburg, Institute of Functional Genomics: Peter Oefner, Wolfram Gronwald, and Department of Medical Biometry, Informatics and Epidemiology (IMBIE), University Hospital of Bonn: Matthias Schmid, Jennifer Nadal.

#### SUPPLEMENTARY METHODS

##### Detailed explanation of filter criteria selection

The GCKD cohort consists of more than 5.000 adult patients with chronic kidney disease of stage 3 or overt proteinuria[1,2] at entry into the registry (eGFR 30 – 60 ml/min). If financial resources were unlimited, it would be preferable to analyse all CKD patients by whole exome sequencing. However, it appears feasible to detect most patients with a hereditary cause of CKD by selecting the most likely probands by distinct criteria and restrict the molecular analysis to the most likely genes.

All participating nephrologists were asked to enter the leading causative reason for CKD, where competing diagnoses were possible. In general, we focused on the diseases which have the potential of being partly or totally misclassified. Usually, hereditary diseases such as ADTKD lead to end stage renal disease (ESRD) latest by 60 years of age. Since most diseases of interest develop over many years if not decades we hypothesize that most patients will have reached CKD stage 3 latest by the age of 50. Therefore, for most categories we have filtered for patients equal or below 50 years of age. The single exception of this rule is the category of “hereditary disease”, where the clinicians will have recognized a positive family history of CKD and thus we did not implement an age restriction. For this group the database provides the defined diagnosis for many distinct diseases, which were excluded for further analysis by individual calling.

Patients with certain diseases where the diagnosis is usually an accurate call, we excluded completely from further analysis. Amongst these were diseases such as ADPKD, microscopic polyangiitis, aHUS/TTP, membranous nephropathy or Lupus erythematodes . We did not exclude patients where diabetes mellitus was entered as the leading cause, since additional development of diabetes is not so unlikely in any patient with a hereditary kidney disease, considering its incidence in the normal population.

Certain diagnoses are in our experience regularly prone to inaccuracy for different reasons. These diagnoses are analgesic nephropathy, chronic glomerulonephritis and IgA nephropathy. In the latter the pathologist may be tempted to discuss IgA, when he sees chronic degenerative histopathology with some amounts of mesangial deposition of IgA. This may in some cases be false, or not the dominant reason for development of CKD. However, since most of these calls will be correct, in these cases we decided to imply more stringency and reduced the age cutoff to 40 years of age, or lower.

### SUPPLEMENTARY RESULTS

#### Additional variants of unknown significance in 9.6%

Further 26 variants of unknown significance (VUS; ACMG class 3) in nine genes (*COL4A3*, *COL4A4*, *COL4A5*, *DNAJB11*, *GATM*, *HNF1B*, *MYH9*, *PARN*, *REN*) were identified in 26 (9.6%) patients (Table 2). As the VUS category has the largest probability span from >10% to <90% chance of being pathogenic, we calculated the odds and p-values of pathogenicity using our manual classification criteria and recently published Bayesian framework.[3] Seven missense variants in seven genes (*COL4A3*, *COL4A4*, *COL4A5*, *DNAJB11*, *GATM*, *MYH9*, *REN*) had an odds-ratio of pathogenicity >9 and a probability of pathogenicity between 0.50 - 0.68. These “hot” class 3 variants could reach the likely pathogenic class with an additional moderate strength criteria like location in an established functional domain (PM1) or if another missense change at the same amino acid residue would be reported as pathogenic (PM5). With automated ACMG classifiers 3/7 (42.9%) of these “hot” VUS would be classified as likely pathogenic by Varsome and all seven (100%) as “VUS/Weak Pathogenic” by VarSeq. Overall, this indicates that further research and stringent deposition of diagnostic variants into public databases could further improve variant classification and thus increase diagnostic yield.

#### Nephronophthisis carrier status is not enriched

Interestingly, we identified 11 carriers (11/271 ~ 4.1%) for (likely) pathogenic variants in six nephronophthisis associated genes (4x *NPHP3*, 2x *CEP290*, 2x *IQCB1*, 1x *ANKS6*, 1x *TTC21B*, 1x *ZNF423*), yet not a single diagnostic case with a second (including VUS) variant *in trans* (Table 3). Thus, nephronophthisis does not appear to play a role in this adult cohort. We asked whether the observed carrier frequency in our cohort represents an enrichment, which could point to either missed deep intronic/ regulatory variants or to nephronophthisis carrier status being a risk factor for CKD. We thus downloaded all variants from the gnomAD database for the 17 nephronophthisis genes in our target design, classified them using the VarSeq classifier and used the allele frequencies for (likely) pathogenic variants detectable by our design to calculate the probability of being a mutation carrier in at least one of these genes (~ 3.5%). As nine variants in our cohort (9/271 ~ 3.3%) would be automatically classified as (likely) pathogenic, the results indicate no enrichment and refutes our initial hypothesis. Knowing this high background carrier probability, reporting of heterozygous carrier status in individuals without a clear clinical suspicion of nephronophthisis should carefully be considered to not cause diagnostic uncertainty in patients and clinicians.

**Comprehensive *MUC1*-VNTR analysis identifies no *MUC1*-dupC**

We performed diagnostic grade SNaPshot mini-sequencing[4] for the typical *MUC1*-dupC variant in the VNTR in all 271 archived DNA samples from the final cohort. For 225 individuals (83.8%) we obtained reliable results but did not identify a *MUC1*-dupC positive case. In 14 samples (5.2%) the results obtained could not reliably be evaluated and in the remaining 32 samples (11.8%) SNaPshot sequencing was not possible, likely due to low DNA quality. Due to the GCKD study design we could not re-contact the individuals/ clinicians to obtain new samples.

We had designed the panel target to directly cover the *MUC1*-VNTR with capture probes and included three known *MUC1*-dupC positive DNA samples from individuals previously diagnosed in our institute.[4,5] Using the adVNTR software, we could confirm the *MUC1*-dupC event (“I22_2_G_LEN1”) in the three positive controls, but did not find this typical duplication or any other high confidence sequence variant in the VNTR, potentially leading to a similar aberrant protein product, in any of the 271 cohort samples.

Thus, combining SNaPshot mini-sequencing and bioinformatic analysis, no ADTKD-*MUC1* case (0/271) could be identified in this cohort of mostly sporadic kidney diseases (Figure 2G). Compare also File S3[6] sheets “SNaPshot” and “adVNTR” for complete per sample results.

#### SUPPLEMENTARY FIGURES

##### Figure S1 | Diagnostic yield and COL4 gene fraction simulations using automated variant classification

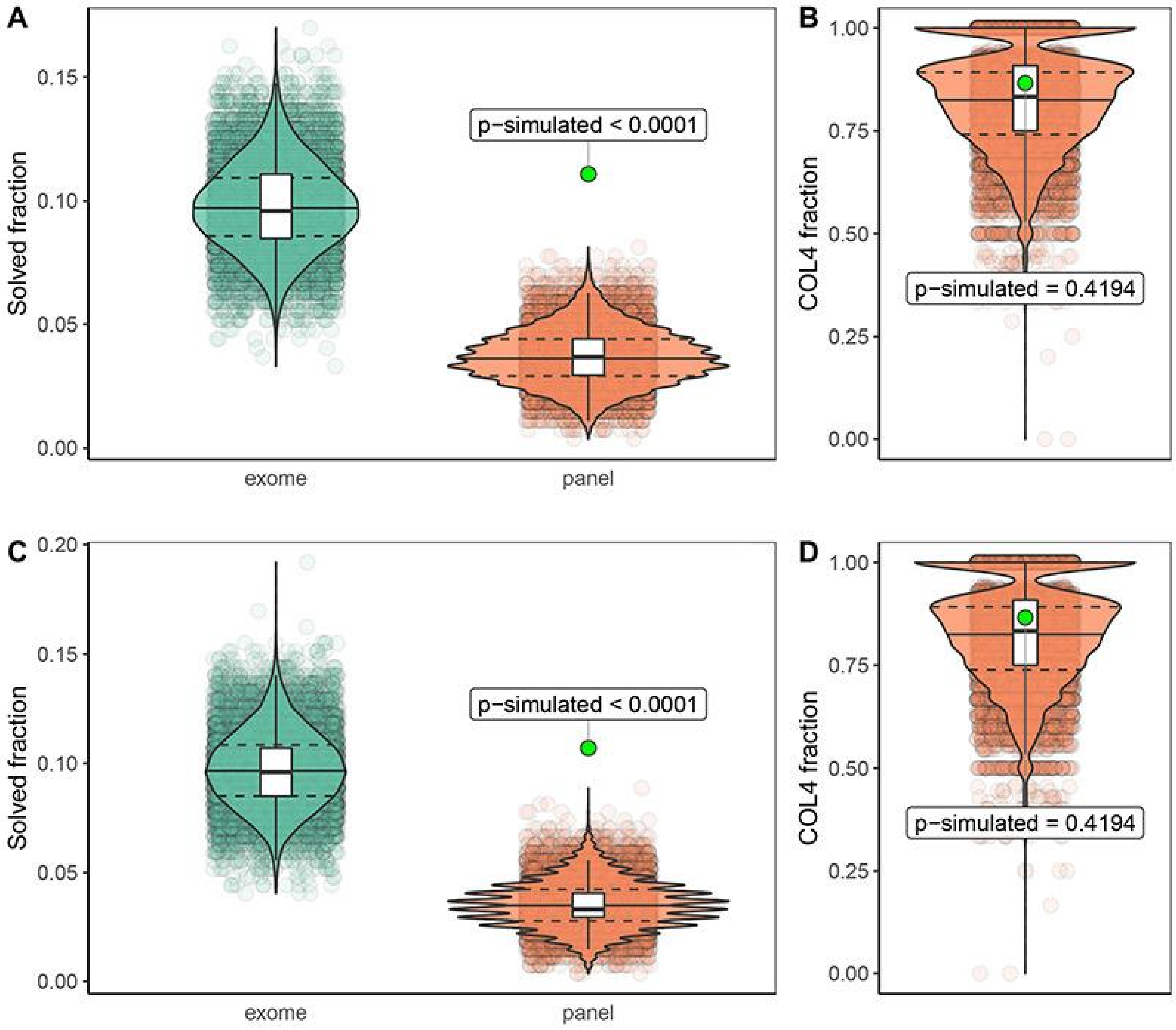


Results as in main Figure 1 but using automated variant classifications for both our and the Groopman cohort variants. **(A)** and **(B)** automated ACMG classification using Varsome. **(C)** and **(D)** automated ACMG classification using VarSeq.

**Figure S2 | Albumin-Creatinine Ratio (ACR) by IgA nephropathy (IgAN) status and genetic diagnosis group**

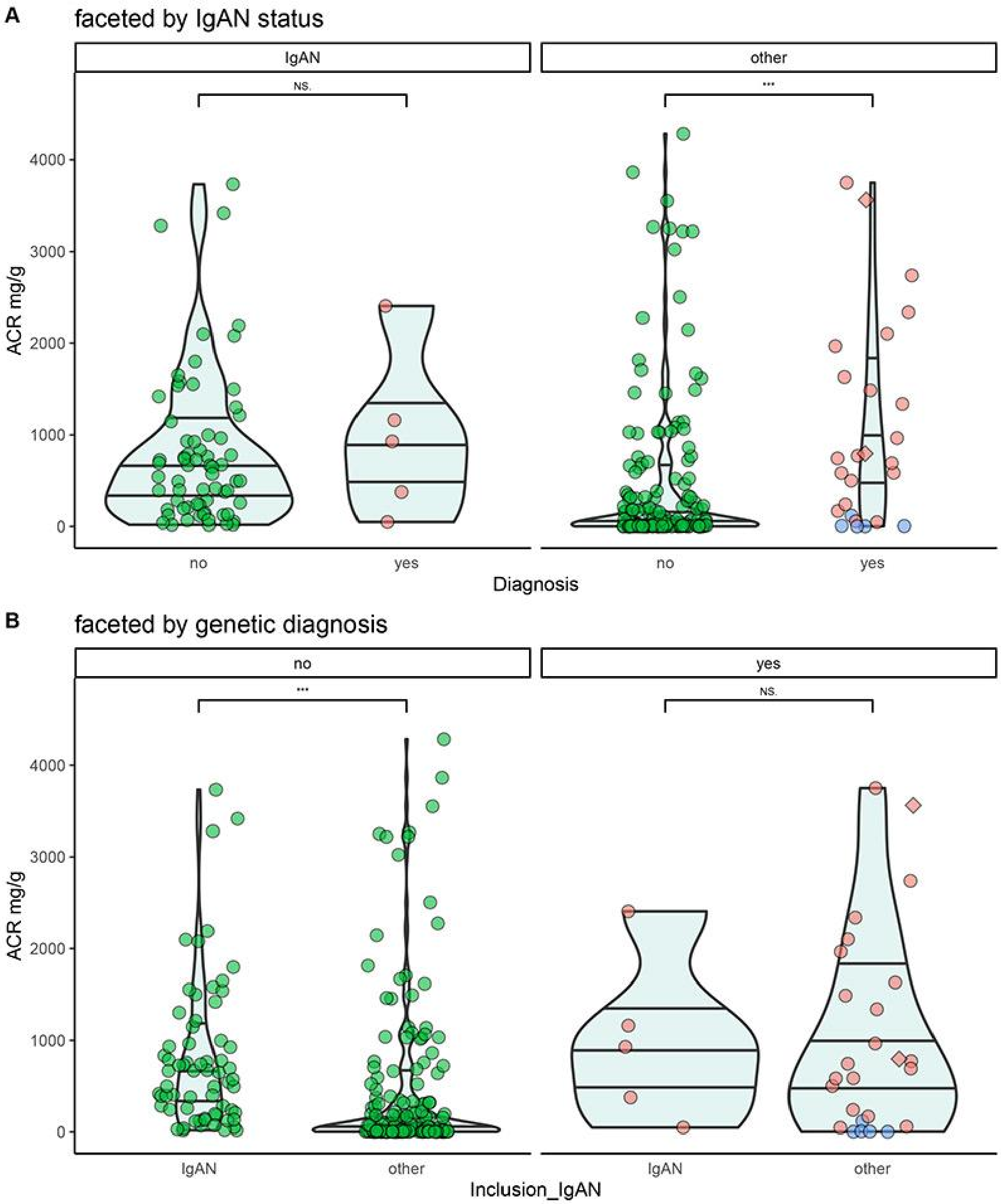


Data on ACR in the cohort from the right panel of Figure 3C faceted by **(A)** IgAN status (left clinical group with IgAN, right other groups) and **(B)** genetic diagnosis group (left no genetic diagnostic variant identified, left with (likely) pathogenic variant). Colored as in Figure 3C. Results indicate that individuals with clinical IgAN have significantly higher ACR, but this does not drive the significantly higher ACR observed in the individuals with a diagnostic variant. This might further support that the clinical classification of IgAN in these individuals with a diagnostic variant is not the primary cause of nephropathy.

#### WEB RESOURCES

gnomAD browser: http://gnomad.broadinstitute.org/

ClinVar: https://www.ncbi.nlm.nih.gov/clinvar/

VariantValidator: https://variantvalidator.org

adVNTR: https://github.com/mehrdadbakhtiari/adVNTR/

RNAfold: http://rna.tbi.univie.ac.at/cgi-bin/RNAWebSuite/RNAfold.cgi

MITOMAP: https://www.mitomap.org/MITOMAP

#### ABBREVIATIONS

ADTKD:autosomal dominant tubulointerstitial kidney disease

CKD: chronic kidney disease

COL4: Collagen-4 genes associated with Alport syndrome (*COL4A5*, *COL4A4* and *COL4A3*)

CNV: copy number variant

DNA: deoxyribonucleic acid

ES: exome sequencing

GCKD: German Chronic Kidney Disease

indel: insertion/ deletion variant

MITKD: mitochondrially inherited tubulointerstitial kidney diseases

NPHP: nephronophthisis

SNV: single nucleotide variant
